# Analysis of COVID-19 and comorbidity co-infection Model with Optimal Control

**DOI:** 10.1101/2020.08.04.20168013

**Authors:** A. Omame, N. Sene, I. Nometa, C. I. Nwakanma, E. U. Nwafor, N. O. Iheonu, D. Okuonghae

**Affiliations:** Department of Mathematics, Federal University of Technology Owerri, Nigeria; Laboratoire Lmdan, Département de Mathématiques de la Décision, Université Cheikh Anta Diop de Dakar, Facultédes Sciences Economiques et Gestion, BP 5683 Dakar Fann, Senegal; Department of Mathematics, University of Hawaii Manoa, USA; Networked Systems Lab, IT Covergence Engineering, School of Electronic Engineering, Kumoh National Institute of Technology Gumi, Korea; Department of Mathematics, University of Benin, Benin City, Nigeria

**Keywords:** COVID-19, Comorbidity, Re-infection, Data-fitting, Optimal Control

## Abstract

The new coronavirus disease 2019 (COVID-19) infection is a double challenge for people infected with comorbidities such as cardiovascular and cerebrovascular diseases and diabetes. Comorbidities have been reported to be risk factors for the complications of COVID-19. In this work, we develop and analyze a mathematical model for the dynamics of COVID-19 infection in order to assess the impacts of prior comorbidity on COVID-19 complications and COVID-19 re-infection. The model is simulated using data relevant to the dynamics of the diseases in Lagos, Nigeria, making predictions for the attainment of peak periods in the presence or absence of comorbidity. The model is shown to undergo the phenomenon of backward bifurcation caused by the parameter accounting for increased susceptibility to COVID-19 infection by comorbid susceptibles as well as the rate of re-infection by those who have recovered from a previous COVID-19 infection. Sensivity analysis of the model when the population of individuals co-infected with COVID-19 and comorbidity is used as response function revealed that the top ranked parameters that drive the dynamics of the co-infection model are the effective contact rate for COVID-19 transmission, *β*_CV_, the parameter accounting for increased sucseptibility to COVID-19 by comorbid susceptibles, *χ*_CM_, the comorbidity development rate, *θ*_CM_, the detection rate for singly infected and co-infected individuals, *η*_1_ and *η*_2_, as well as the recovery rate from COVID-19 for co-infected individuals, *φ*_I2_. Simulations of the model reveal that the cumulative confirmed cases (without comorbidity) may get up to 180,000 after 200 days, if the hyper susceptibility rate of comorbid susceptibles is as high as 1.2 per day. Also, the cumulative confirmed cases (including those co-infected with comorbidity) may be as high as 1000,000 cases by the end of November, 2020 if the re-infection rates for COVID-19 is 0.1 per day. It may be worse than this if the re-infection rates increase higher. Moreover, if policies are strictly put in place to step down the probability of COVID-19 infection by comorbid susceptibles to as low as 0.4 per day and step up the detection rate for singly infected individuals to 0.7 per day, then the reproduction number can be brought very low below one, and COVID-19 infection eliminated from the population. In addition, optimal control and cost-effectiveness analysis of the model reveal that the the strategy that prevents COVID-19 infection by comorbid susceptibles has the least ICER and is the most cost-effective of all the control strategies for the prevention of COVID-19.

## 1 Introduction

The new coronavirus disease 2019 (COVID-19) was reported for the first time in the Chinese city of Wuhan, in the closing month of the year 2019 [22]. The disease spread rapidly across the globe like wildfire, that on March 11, 2020, the World Health Organization (WHO) delared it a global pandemic [11]. As of 25th July, 2020, the global cases of COVID-19 infection stands at 15,762,007 with 640,276 deaths [12]. The morbidity and mortality vary across countries of the world with the highest reported in the United States (4,113,224 confirmed cases with 145,556 deaths), and Brazil (2,287,475 confirmed cases with 85,238 deaths) as of 25th July, 2020. [12]. COVID-19 “can often present as a common cold-like illness”, with other symptoms including fever, fatigue, muscle pains, loss or change of taste or smell, shortness of breath, dry cough and sore throat [47].

The disease can be spread from person-to-person through the breathing in of respiratory droplets from infected persons and via direct contact with contaminated objects and surfaces [2]. Although no vaccine or antiviral treatments have been approved for the prevention or management of COVID-19, governments of different nations and individuals have embarked on non-pharmaceutical interventions such as face-mask usage in public places, social disancing observance and lockdown imposition as a means of effectively reducing the spread of the disease [4, 42]. Epidemiological studies have recently pointed towards the need to consider COVID-19 and comorbidity (such as diabetes, lung disease and heart disease) co-infection, as those with the latter have a higher chance of infection with the former [11]. Bjorgul *et al*. [6] defined comorbidities as “diseases or medical conditions unrelated in etiology or causality to the principal diagnosis that coexist with the disease of interest”. According to the Centres for Disease Control and prevention (CDC), “People of any age with the following conditions are at increased risk of severe illness from COVID-19: Chronic kidney disease, COPD (chronic obstructive pulmonary disease), Immunocompromised state (weakened immune system) from solid organ transplant, Obesity (body mass index [BMI] of 30 or higher), Serious heart conditions, such as heart failure, coronary artery disease, or cardiomyopathies, Sickle cell disease, Type 2 diabetes mellitus, Hypertension or high blood pressure and Neurologic conditions, such as dementia” [10]. This is further confirmed by a clinical report by Huang *et al*. [18] who in a survey of 41 confirmed COVID-19 patients discovered that thirteen of them had underlying diseases, which include, hypertension, chronic obstructive pulmonary disease cardiovascular disease and diabetes. Furthermore, Wang *et al*. reported from a finding incolving 138 patients, that over 45% of them had one or more comorbidities and that “the patients who were admitted to the intensive care unit (ICU) had a higher number of comorbidities (72.2%) than those not admitted to the ICU (37.3%)” [46, 49]. In Nigeria, the Ministry of Health had reported that prior comorbid conditions such as diabetes mellitus, tuberculosis and hypertension were responsible for about 70% of COVID-19 related deaths [38]. Recent studies carried out in Nigeria estimated the prevalence of diabetes mellitus (DM) to range from lowest level of 0.8% among adults in rural settings to over 7% in urban Lagos with an average of 2.2% nationally [15]. Diabetes mellitus has been reported to increase the vulnerability of individuals to infection with COVID-19 in Nigeria. The prevalence of DM is higher in patients with severe COVID-19 disease than in patients with mild symptoms [33]. More recently, WHO has warned that currently no evidence to show that those who have recovered from COVID-19 infection can not get re-infected [48]. On April 13, 2020, South Korea reported that more than 100 of its recovered cases of COVID-19 have been re-infected again [41]. This calls for serious concern as several countries, including China and South Korea, which had previously recorded giant strides in curbing the the virus, have recently reported second waves of COVID-19 infections [5, 14].

Mathematical modelling has been a powerful means of studying the behaviour of infectious diseases [13, 30, 31, 34, 35, 36, 40, 39, 43]. A lot of models have been developed for the dynamics of COVID-19 [1, 16, 19, 20, 21, 28, 32]. In this study, we shall be investigating the impact of re-infection and coinfection with comorbidity on the dynamics of COVID-19 disease. In addition, we shall be carrying out optimal control analysis on the model to assess the impact of some control strategies on the prevention of COVID-19. Specifically, the comorbidity of interest for this study shall be **diabetes mellitus**.

The paper is organized as follows. The model formulation and basic properties of the model are reported in Section 2. The model (without controls) is analyzed (qualitatively and quantitatively) in Sections 3 and 4. Optimal control analysis is carried out in Section 5 while Section 6 gives the concluding remarks.

## 2 Model formulation

The total population at time *t*, denoted by *N*_H_(*t*), is divided into eight mutually-exclusive compartments: Susceptible individuals (*S*_H_(*t*)), comorbid susceptible individuals (*S*_CM_), individuals with COVID-19 infection (*I*_CV_(*t*)), isolated and hospitalized individuals infected with COVID-19 (*Q*_CV_(*t*)), individuals who have recovered from COVID-19 infection (*R*_CV_(*t*)), individuals co-infected with COVID-19 and comorbidity (*I*_OVOM_(*t*)), isolated and hospitalized individuals co-infected with COVID-19 infection and comorbidity (*Q*_CVCM_), individuals who have recovered from COVID-19 infection but with comorbidity (*R*_CM_). Susceptible individuals acquire COVID-19 infection at the rate

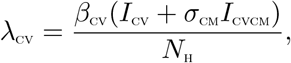

where *σ*_CM_ accounts for increased infectiousness of co-infected individuals. Furthermore, susceptible individuals also acquire comorbidity at the rate *θ*_CM_. Comorbid susceptible individuals acquire COVID-19 infection at an increased rate *χ*_CM_*λ*_CV_, with *χ*_CM_ > 1. Natural death occurs in all compartments at the rate *μ*_H_. It is imperative to state at the onset that the comorbidity considered in this study is **diabetes mellitus**.

The total population at time, t is given by

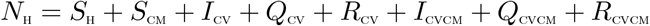

Based on the above formulations and assumptions, the COVID-19 and comorbidity co-infection model is given by the system of non-linear deterministic differential equations (flow diagram of the model is shown in Figure 1, the associated state variables and parameters are well described in Table 1) given below:

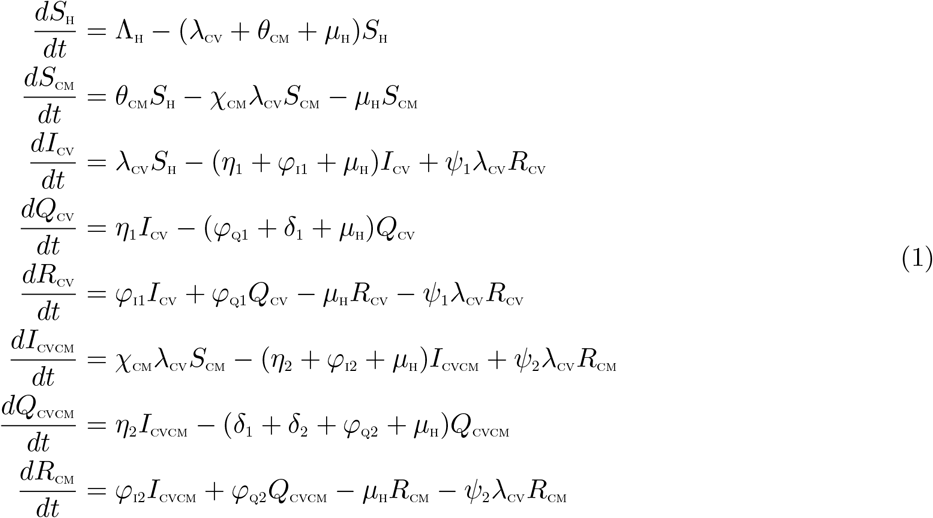

with

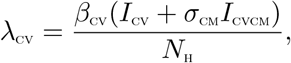

**Table 1:**
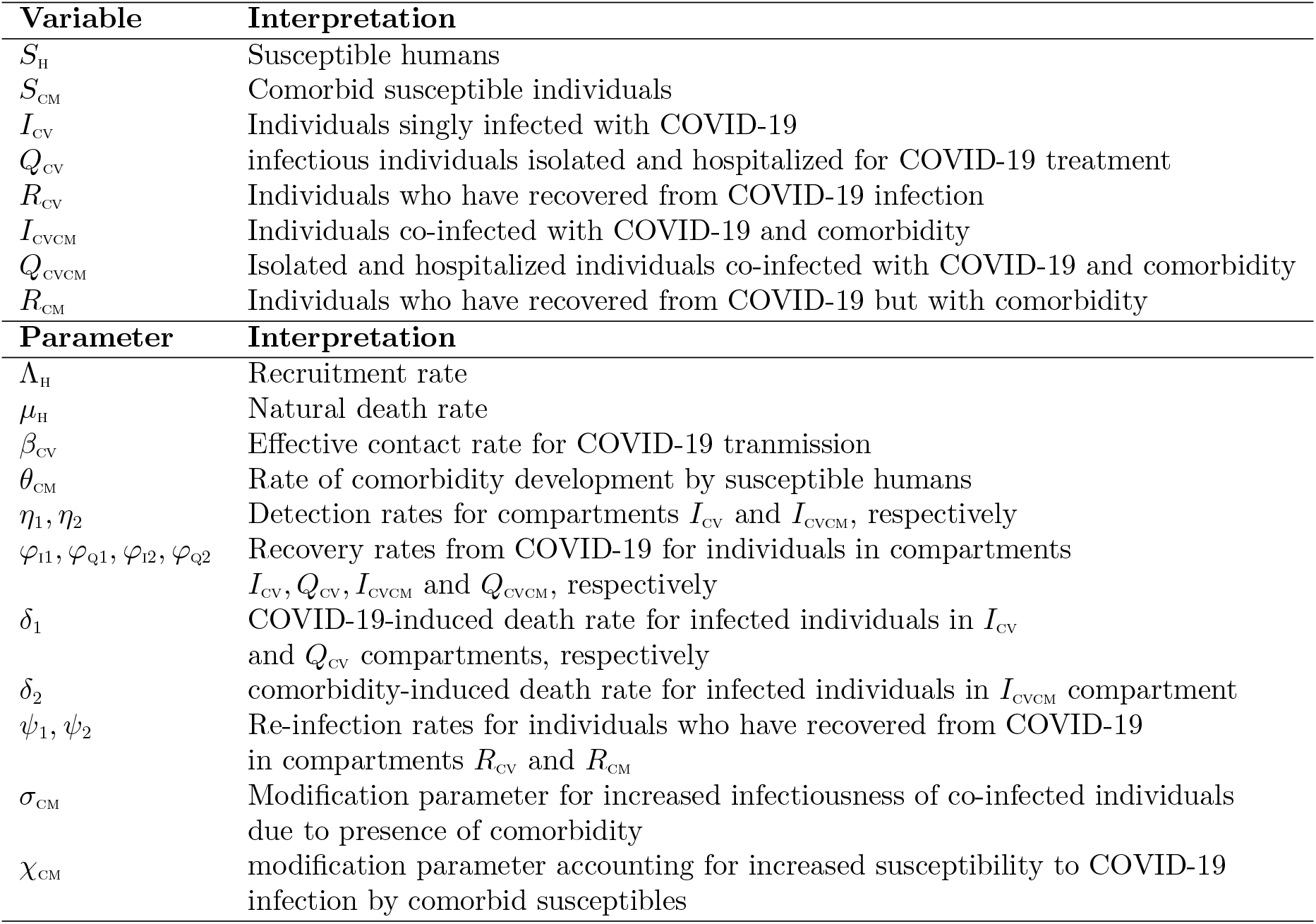
Description of variables and parameters in the model equation

**Figure 1:**
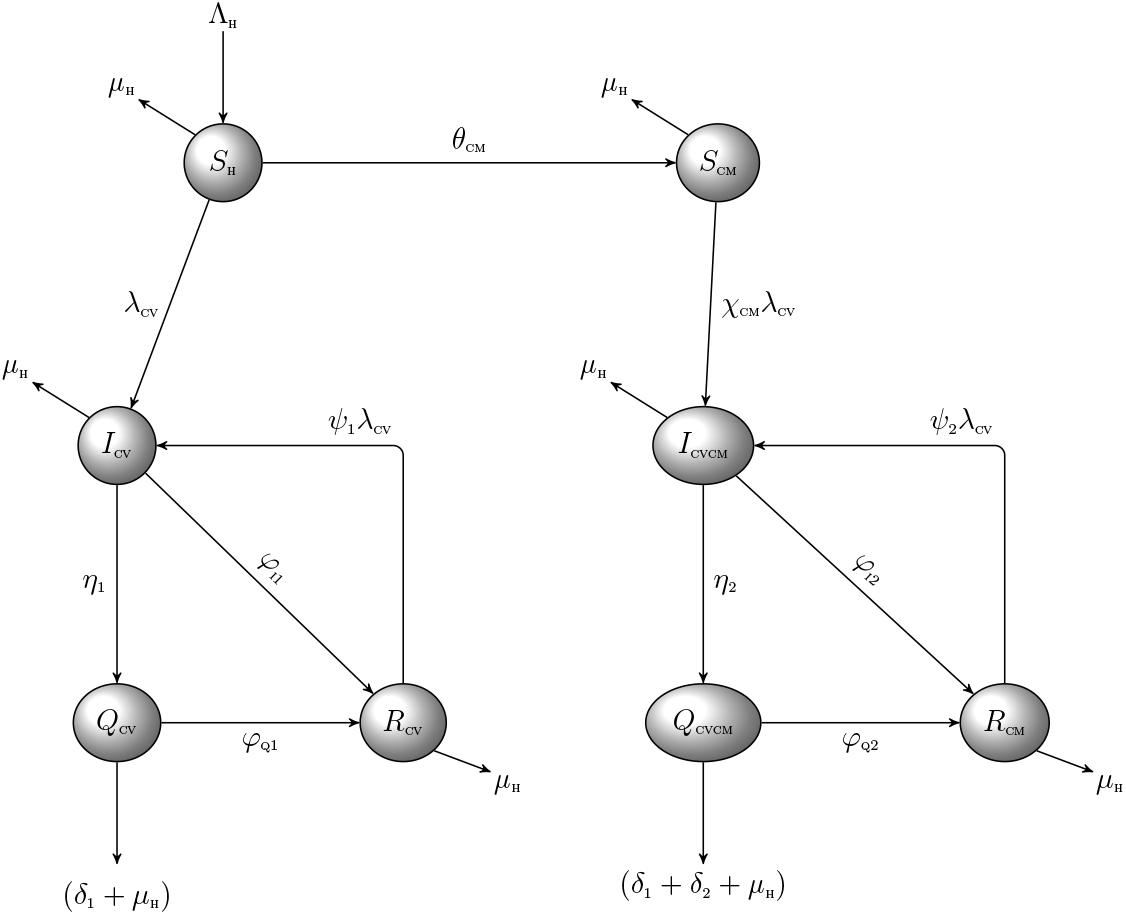
Schematic diagram of the model (1) where 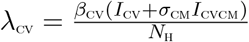

### 2.1 Basic properties of the model

#### 2.1.1 Positivity and boundedness

For the model (1) to be epidemiologically meaningful, it is important to prove that all its state variables are non-negative for all time (*t*).

##### Theorem 2.1

*Let the initial data S*_H_ *>* 0, *S*_CM_ *>* 0, *I*_CV_ *>* 0,*Q*_cv_ *>* 0, *R*_cv_ *>* 0*; I*_CVCM_ *>* 0,*Q*_CVCM_ *>* 0*;R*_CM_ *>* 0. *Then the solutions* (*S*_H_, *S*_CM_, *I*_CV_, *Q*_cv_, *R*_cv_, *I*_CVCM,_ *Q*_CVCM_*, R*_CM_) *of the model* (1) *are positive for all time t >* 0.

###### Proof.

Let

*t*_1_ = sup{*t* > 0: *S*_H_ > 0*, S*_CM_ > 0*, I*_CV_ > 0*,Q*_CV_ > 0*,R*_CV_ > 0*, I*_CVCM_ > 0*,Q*_CVCM_ > 0*,R*_CM_ > 0 ∈ [0*, t*]}. Thus, *t*_1_ *>* 0.

We have, from the first equation of the system (1) that

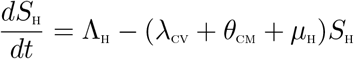

which can be re-written as

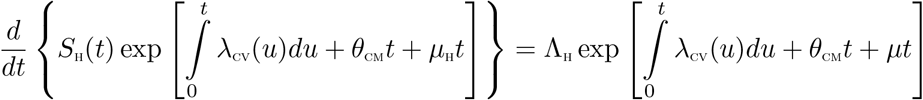

so that

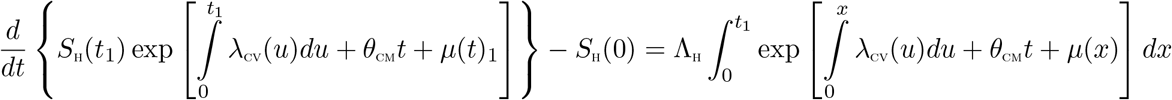

so that

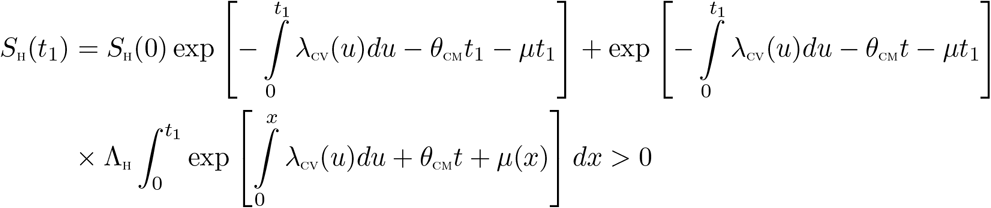

In a similar manner, it can be proven that:

*S*_CM_ > 0, *I*_cv_ > 0, *Q*_cv_ > 0, *R*_CV_ > 0, *I*_CVCM_ > 0, *Q*_CVCM_ > 0, *R*_CM_ > 0

### 2.2 Invariant regions

The co-infection model (1) will be analyzed in a biologically feasible region as follows. We first show that the system (1) is dissipative in a proper subset 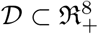. Let

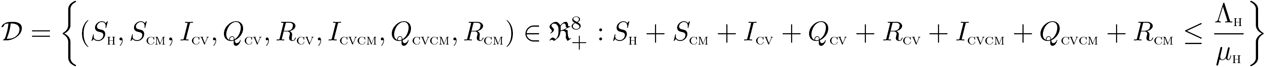

The following steps are followed to establish the positive invariance of 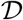.

Adding all the equations of the system (1) gives

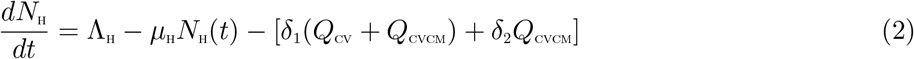

From (2), we have that

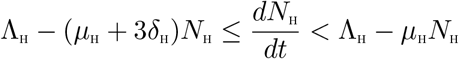

where *δ*_H_ = min{*δ*_1_, *δ*_2_}

Using the Comparison theorem [23], we have that, 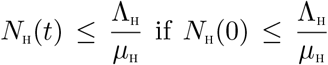. Thus, the region 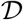 is positively invariant. Hence, it is sufficient to consider the dynamics of the flow generated by the system (1) in 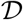. Thus, within this region, the model (1) is said to be be epidemiologically and mathematically well-posed [17]. Thus, every solution of the model (1) with initial conditions in 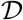 remains in 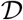 for all time *t* ≥ 0. Therefore, the *ω*—limit sets of the system (1) are contained in *D*. This result is summarized thus.

#### Lemma 2.1

*The region* 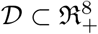 *is positively-invariant for the model* (1) *with initial conditions in* 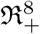.

## 3 Analysis of the model without controls

In this section, we seek to qualitatively study the dynamical properties of the model (1) without controls.

### 3.1 Basic reproduction number of the co-infection model (1)

The COVID-19-comorbidity co-infection model (1) has a COVID-19 free equilibrium (CFE), obtained by setting the disease classes and the right-hand sides of the equations in the model (1) to zero, given by

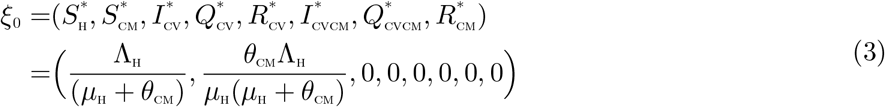

The basic reproduction number of the co-infection model (1), using the approach illustrated in [44], is given by

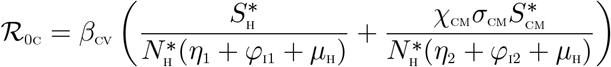

### 3.2 Local asymptotic stability of the COVID-19 Free Equilibrium (CFE) of the coinfection model

#### Theorem 3.1

*The CFE, ξ*_0_, *of the model* (1) *is locally asymptotically stable (LAS*) *if* 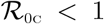*, and unstable if* 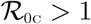.

##### Proof.

The local stability of the model (1) is analysed by the Jacobian matrix of the system (1) evaluated at the COVID-19-free equilibrium *ξ*_0_, given by:

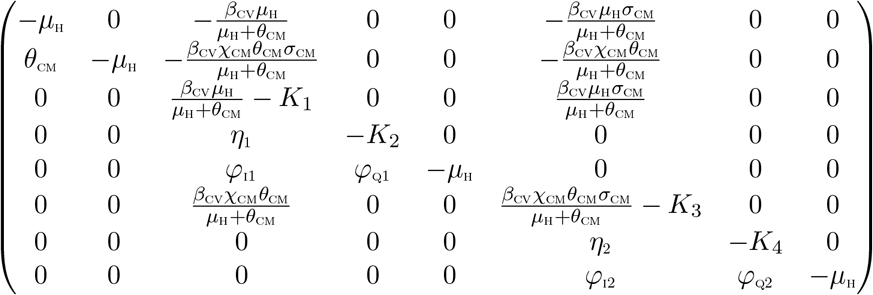

where, *K*_1_ = *η*_1_ + *φ*_I1_ + *μ*_H_, *K*_2_ = *φ*_Q1_ + *δ*_1_ + *μ*_H_, *K*_3_ = *η*_2_ + *φ*_I2_ + *μ*_H_, + *K*_4_ = *φ*_Q2_ + *δ*_1_ + *δ*_2_ *+ μ*_H_.

The eigenvalues are given by *λ*_1_ = −(*η*_1_ + *φ*_I1_ + *μ*_H_), *λ*_2_ = −(*φ*_Q1_ + *δ*_1_ + *μ*_H_), *λ*_3_ = −(*η*_2_ + *φ*_I2_ + *μ*_H_), *λ*_4_ = −(*φ_Q_*_1_ + *δ*_1_ + *δ*_2_ + *μ*_H_)

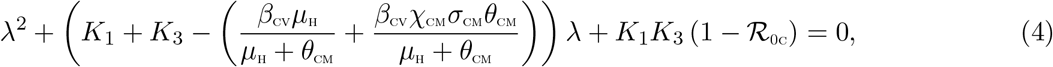

Applying the Routh-Hurwitz criterion, the quadratic equation (4) will have roots with negative real parts if and only if 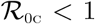. As a result, the COVID-19-free equilibrium, *ξ*_0_ is locally asymptotically stable if 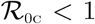.

The epidemiological implication of Theorem 3.1 is that COVID-19 can be eliminated from the population when 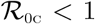 and if the initial sizes of the population of the model are in the region of attraction of the CFE.

### 3.3 Existence of Endemic equilibrium points (EEP) of the model

In this section, we shall investigate the existence of an endemic equilibrium of the model (1). Let an arbitrary equilibrium point of the model be given by

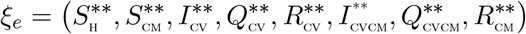

The steady state solutions of equations of the model (1)

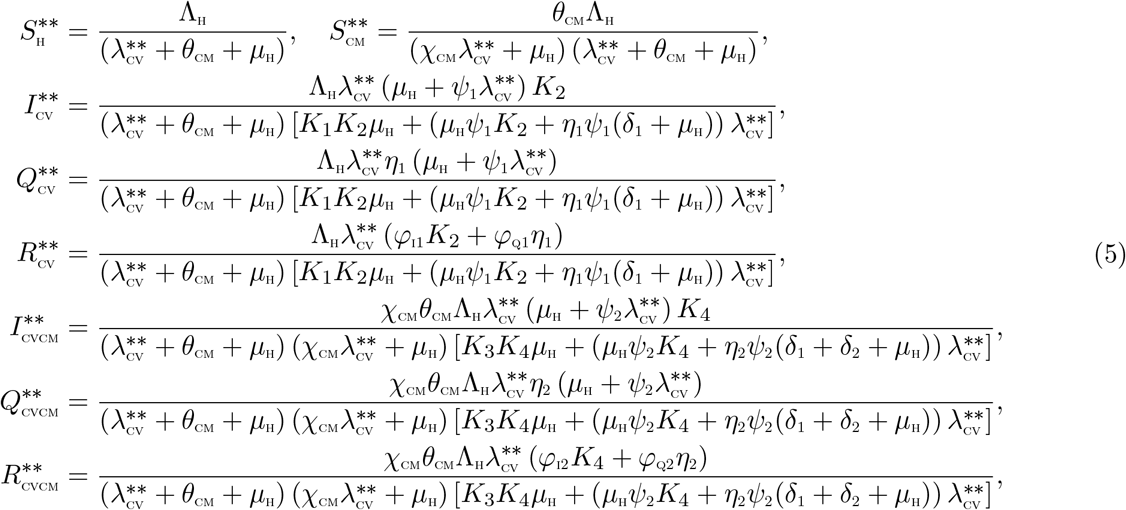

Substituting the above expressions into the force of infection, at steady state, gives the following polynomial

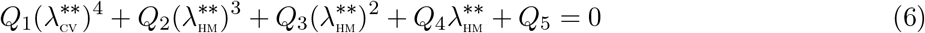

where

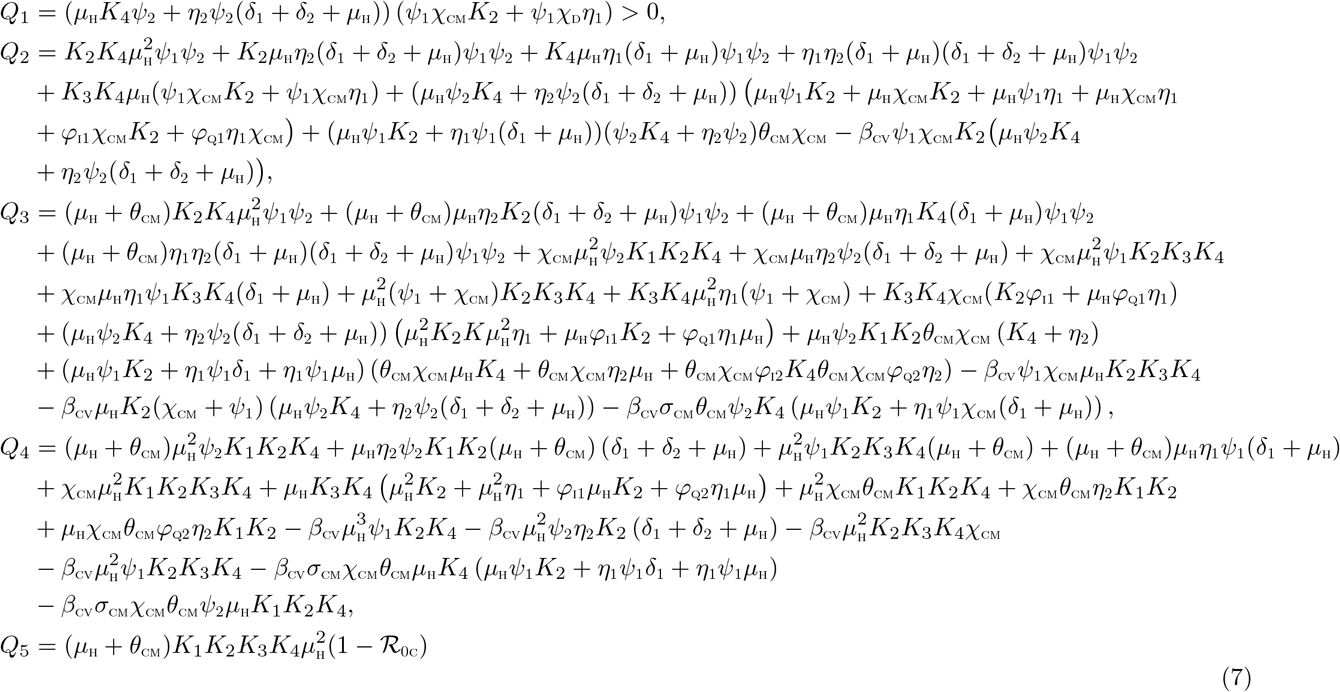

The components of the EEP are obtained upon solving for 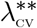 from the polynomial (6), and substituting the positive values of 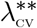 into the expressions in (5). Moreover, it follows from (7) that the coefficient *Q*_1_ is always positive, and *Q*_5_ is positive (negative) if 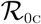 is less (greater) than one. The following results can be deduced.

#### Theorem 3.2

The *model* (1) *has:*

i. *four or two endemic equilibria if Q*_2_ > 0, *Q*_3_ < 0, *Q*_4_ > 0 *and* 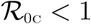,
ii. *two endemic equilibria if Q*_2_ > 0, *Q*_3_ > 0, *Q*_4_ < 0 *and* 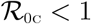
iii. *no endemic equilibrium otherwise, if* 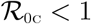,

The first two items of Theorem 3.2 ((i) - (ii)) suggest the possibility of backward bifurcation in the model (1) when 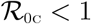. It is worthy of note to show that, setting the parameters *ψ*_1_ = *ψ*_2_ = *χ*_CM_ = 0, reduces the quartic (6) to 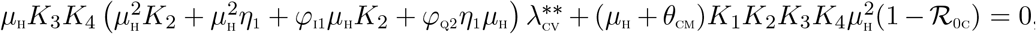, resulting in no sign changes in the polynomial equation for 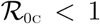. However, for 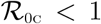 a unique endemic equilibrium exists. In the subsequent section, we shall explore the existence of the phenomenon of backward bifurcation in the model (1).

### 3.4 Backward bifurcation analysis of the model (1)

In this section, we shall seek to determine the type of bifurcation the model (1) will exhibit, using the Center Manifold theory presented by Castillo-Chavez and Song [8]. We establish the result below

#### Theorem 3.3

*Suppose a backward bifurcation coefficient a* > 0, *(with a defined below*)*, when* 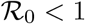

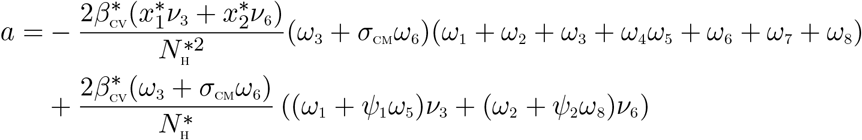

*then model* (1) *undergoes the phenomenon of backward bifurcation at* 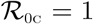. *If a* < 0, *then the system* (1) *exhibits a forward bifurcation at* 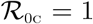.

##### Proof

Suppose

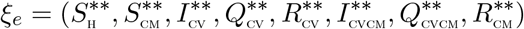

represents any arbitrary endemic equilibrium of the model. To apply the Center Manifold theory, it is important we carry out the change of variables below.

Let

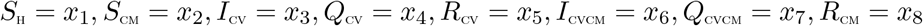

Moreover, using the vector notation

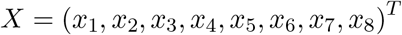

the model (1) can be re-written in the form

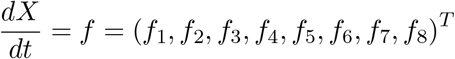

as follows:

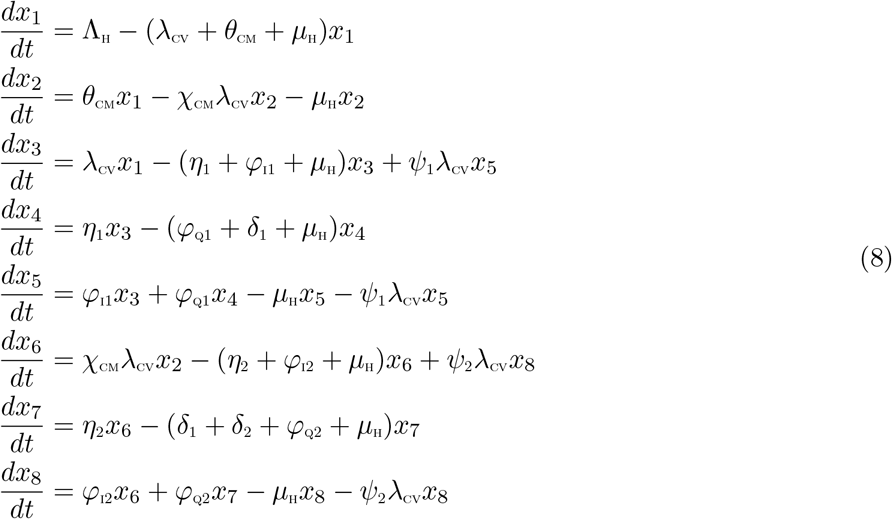

**Figure 2:**
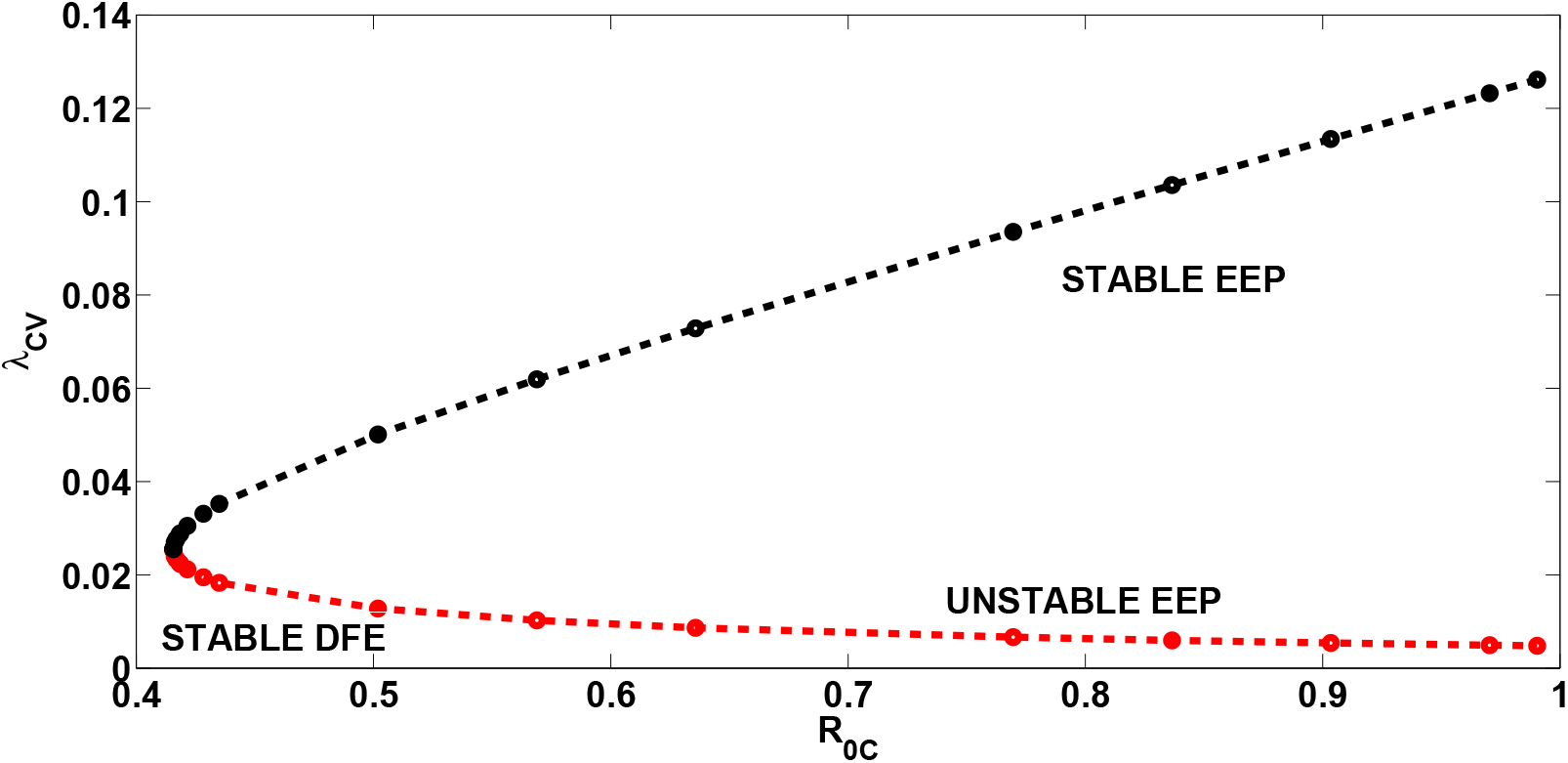
Bifurcation diagram for the model (1). Parameter values used are: *β*_CV_ = 0.148, *ψ*_1_ = 25, *ψ*_2_ = 40, *χ*_CM_ = 1.2. All other parameters as in Table 1

with

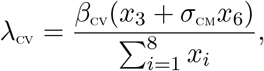

Consider the case when 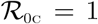. Moreover, let *β*_CV_ be chosen as a bifurcation parameter. Solving for 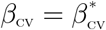 from 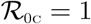 we obtain

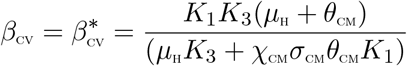

Evaluating the Jacobian of the system (1) at the CFE, *J*(*ξ*_0_), and evaluating the right eigenvector, **w** = [*ω*_1_ *ω*_2_, *ω*_3_, *ω*_4_, *ω*_5_, *ω*_6_, *ω*_7_, *ω*_8_]*^T^*, associated with the simple zero eigenvalue of *J*(*ξ*_0_), gives

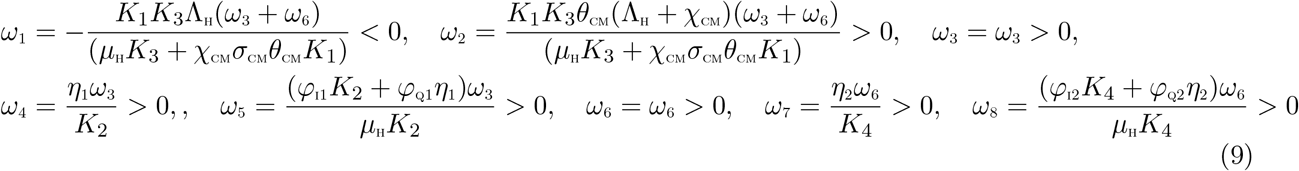

Likewise, the components of the left eigenvector of 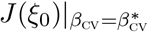, **v** = (*v*_1_, *v*_2_,…,*V*_8_), satisfying **v**.**w** = 1 are

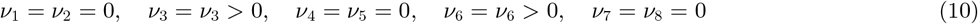

The associated bifurcation coefficients defined by a and b, given by:

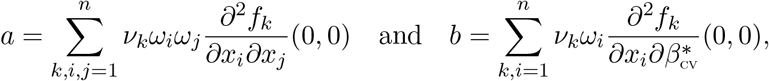

are computed to be

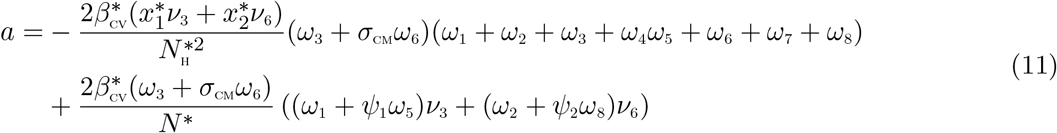

and

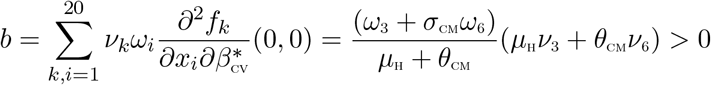

It is observed that the bifurcation coefficient *b* is positive. Hence, following from Theorem 4.1 in [8] we have that the model (1) exhibits backward bifurcation phenomenon if the backward bifurcation coefficient, *a*, given by (11) is positive. The associated backward bifurcation diagram is presented in Figure 2. It is imperative to note, that setting the COVID-19 re-infection terms *ψ*_1_ = *ψ*_2_ = 0 as well as the modification parameter for increased susceptibility to COVID-19 by comorbid susceptibles *χ*_CM_ = 0, we observed that the bifurcation coefficient, *a* < 0. As a result, backward bifurcation does not occur in the COVID-19- comorbidity co-infection model, when there is no re-infection after recovery from COVID-19 and when appropriate measures are enforced to prevent comorbid susceptible individuals from getting infected with COVID-19. This result is consistent with the result obtained in Section 3.3 above. The epidemiological interpretation is that if this phenomenon occurs, then the control of COVID-19 at the community level becomes difficult, even when the associated reproduction number 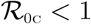.

### 3.5 Global asymptotic stability of the CFE of the model

By removing the cause of the backward bifurcation in the model (1), that is, setting *ψ*_1_ = *ψ*_2_ = *χ*_CM_ = 0, we have the following reduced model.

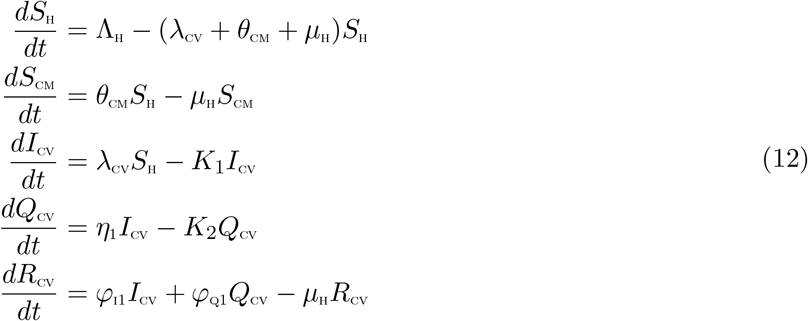

with

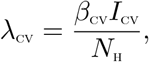

We can prove the global asymptotic stability of the CFE of the model (1).

#### Theorem 3.4

*Consider the model* (1) *with ψ*_1_ = *ψ*_2_ = *χ*_CM_ = 0. *The CFE is GAS in 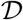 whenever 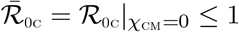*

##### Proof.

We notice the compartments *S*_CM_, *Q*_CV_ and *R*_CV_, do not impact the dynamics of COVID-19 in the considered epidemic model. Therefore, we continue our proof with the compartments *S*_H_, and *I*_cv_. For the rest of the proof, we consider the following Lyapunov function

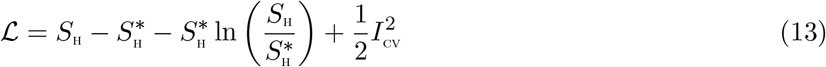

Applying the derivative of the Lyapunov function (13) along the trajectories of the considered model, we have the following relationships

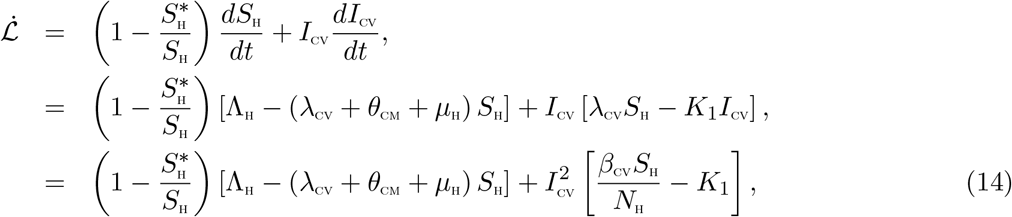

At the COVID-19 free equilibrium point (CFE), we have the following form

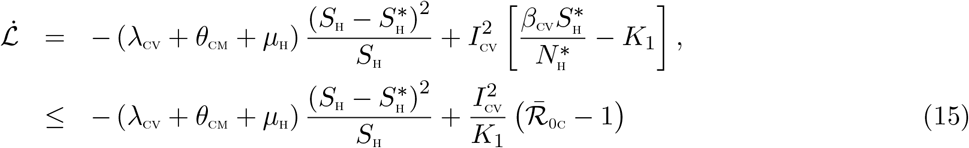

Hence, 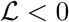 if and only if 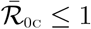. Therefore, 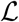 is a Lyapunov function for the system (12). It follows by the La Salle’s Invariance Principle [24], that the CFE of the model (12) is globally asymptotically stable whenever 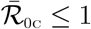.

## 4 Numerical simulations

### 4.1 Uncertainty and sensitivity analyses

As a result of the uncertainties which are expected to come up in parameter estimates used in the numerical simulations, a Latin Hypercube Sampling (LHS) [3] is implemented on the parameters of the model. For the sensitivity analysis, we carry out a Partial Rank Correlation Coefficient (PRCC) between values of the parameters in the response function and the values of the response function derived from the sensitivity analysis. 1,000 simulations of the co-infection model (1) *per* LHS were run. Using the reproduction number, 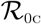, as the response function, we observed in Table 2 that the top-ranked parameters that drive the dynamics of the co-infection model are the effective contact rate for COVID-19 transmission, *β*_CV_, the parameter accounting for increased for susceptibility to COVID-19 by comorbid susceptible individuals, *χ*_CM_, the parameter accounting for the infectiousness of co-infected individuals, *σ*_CM_, the detection rates for singly infected and co-infected individuals, *η*_1_ and *η*_2_, respectively. Also, when the total number of individuals singly infected with COVID-19, *I*_cv_, is used as the response function, the top ranked parametrs that influnce the dynamics of COVID-19 disease are the effective contact rate for COVID-19 transmission, *β*_cv_, the parameter accounting for increased for the infectiousness of co-infected individuals, *σ*_CM_, the detection rates for singly infected and co-infected individuals, *η*_1_ and *η*_2_, as well as the recovery rate from COVID-19, *φ*_11_.

Using the total number of isolated/hospitalized individuals with COVID-19, (*Q*_CV_), as the response function, the parameters that strongly determine the dynamics of the model (1) are the effective contact rate for COVID-19 transmission, *β*_cv_, the parameter accounting for increased for the infectiousness of co-infected individuals, *σ*_CM_, the detection rates for singly infected and co-infected individuals, *η*_1_ and *η*_2_, as well as the recovery rates from COVID-19, *φ*_I1_ and *φ*_Q1_ for individuals in compartments *I*_cv_ and *Q*_cv_, respectively. In addition, when the population of individuals co-infected with COVID-19 and comorbidity is used as response function, the top ranked parameters that drive the dynamics of the co-infection model are the effective contact rate for COVID-19 transmission, *β*_cv_, the parameter accounting for increased sucseptibility to COVID-19 by comorbid susceptibles, *χ*_CM_, the comorbidity development rate, *θ*_CM_, the detection rate for singly infected and co-infected individuals, *η*_1_ and *η*_2_, as well as the recovery rate from COVID-19 for co-infected individuals, *φ*_I2_. Moreover, when the population of isolated/hospitalized individuals with COVID-19 and comorbidity *Q*_CVCM_, is used as response function, the parameters that drive the dynamics of the co-infection model are the effective contact rate for COVID-19 transmission, *χ*_CM_, the parameter accounting for increased sucseptibility to COVID-19 by comorbid susceptibles, *χ*_CM_, the comorbidity development rate, *θ*_CM_, the detection rates for co-infected individuals, *η*_2_, as well as the recovery rates from COVID-19, *φ*_I2_ and *φ*_Q2_ for individuals in compartments *I*_cv_ and *Q*_cv_, respectively.

### 4.2 Model Fitting and estimation of parameters and initial conditions

The population of Lagos is approximately estimated at 14,368,332 [45]. Also, based on a study carried out by Dahiru *et al*. [15] on the prevalence of diabetes mellitus in Lagos, Nigeria and since the first COVID-19 case in Lagos (with no evidence of comorbidity) was announced on March 16, 2020, we set our initial conditions to be *S*_H_(0) = 0.93 × 14368332, *S*_CM_(0) = 0.07 × 14368332, *Q*_cv_(0) = 1, *R*_CV_(0) = 0, *Q*_CVCM_ = 0, *R*_CM_(0) = 0.

Two optimization algorithms were combined for data fitting, a genetic algorithm (GA) algorithm and the *fmincon* algorithm [27, 32] in MATLAB to estimate the values of the effective contact rate for COVID-19 transmission, *β*_cv_, the case detection rates for singly infected and co-infected individuals, *η*_1_ and *η*_2_, respectively, the comorbidity induced death rate for co-infected individuals, *δ*_2_ and the COVID-19 reinfection rates *ψ*_1_ and *ψ*_2_ for singly and co-infected individuals respectively. We equally estimated the initial conditions *I*_CV_(0) and *I*_CVCM_(0). All the estimates were done using the daily confirmed and active cases for Lagos, Nigeria, from March 16, 2020 to June 24, 2020 [29]. Table 3 gives the estimated values of *β*_CV_,*η*_1_,*η*_2_,*δ*_2_,*ψ*_1_ and *ψ*_2_, and the initial conditions *I*_CV_(0) and *I*_CVCM_(0), together with the calculated reproduction number (*R*_0C_).

**Table 2:** PRCC values for the co-infection model (1) parameters using the total number of infected individuals and the reproduction number 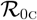 as response functions.

| Parameters | $I_{CV}$ | $Q_{CV}$ | $I_{CVCm}$ | $Q_{CVCm}$ | $\mathcal{R}_{0c}$ |
| --- | --- | --- | --- | --- | --- |
| $\mu_H$ | -0.0856 | -0.0287 | -0.0153 | -0.0212 | -0.0085 |
| $\beta_{CV}$ | **0.9800** | **0.9649** | **0.8857** | **0.7429** | **0.9348** |
| $\chi_{CM}$ | 0.2080 | 0.1715 | **0.5413** | **0.4001** | **0.6883** |
| $\theta_{CM}$ | 0.0989 | 0.1022 | **0.4531** | **0.4031** | -0.0027 |
| $\sigma_{CM}$ | **0.6956** | **0.6280** | 0.3828 | 0.2436 | **0.6079** |
| $\eta_1$ | **-0.9456** | **0.6872** | **-0.5373** | -0.3472 | **-0.6444** |
| $\eta_2$ | **-0.6434** | **-0.5263** | **-0.9000** | **0.7933** | **-0.7336** |
| $\varphi_{i1}$ | **-0.6817** | **-0.5669** | -0.2234 | -0.0913 | -0.2770 |
| $\varphi_{i2}$ | -0.2951 | 0.2156 | **-0.5231** | **-0.4052** | -0.3436 |
| $\varphi_{q1}$ | 0.0112 | **-0.7534** | -0.0309 | 0.0457 | — |
| $\varphi_{q2}$ | 0.0173 | 0.0393 | 0.0738 | **-0.9090** | — |
| $\delta_1$ | 0.0346 | -0.2450 | 0.0087 | -0.3801 | — |
| $\delta_2$ | 0.0110 | -0.0187 | 0.0119 | -0.3830 | — |
| $\psi_1$ | -0.0065 | 0.0512 | -0.0839 | -0.0315 | — |
| $\psi_2$ | 0.0184 | 0.0034 | 0.0522 | 0.0365 | — |

**Table 3:** Estimated parameters and variables when the model 1 was fitted using the cumulative number of confirmed and active cases. The estimates were from the “best fit”.

| Parameter | Baseline value | Range | Reference |
| --- | --- | --- | --- |
| $\mu_H$ | 0.0000491 /day | | [26] |
| $\Lambda_H$ | 705 /day | | [45, 26] |
| $\beta_{CV}$ | 0.2001 | [0.1, 0.3] | Fitted |
| $\varphi_{i1}, \varphi_{i2}$ | 0.13978 /day | $[\frac{1}{30}, \frac{1}{3}]$/day | [42] |
| $\varphi_{q1}, \varphi_{q2}$ | $\frac{1}{15}$ /day | $[\frac{1}{30}, \frac{1}{3}]$/day | estimated from [9] |
| $\delta_1$ | 0.015 /day | [0.001, 0.1] | [16] |
| $\eta_1$ | 0.0059 /day | [0.001, 0.05] | Fitted |
| $\eta_2$ | 0.0895 /day | [0.001, 0.05] | Fitted |
| $\theta_{CM}$ | 0.0000034294 | [0.0000001, 0.00001] | Fitted |
| $\psi_1$ | 0.0013 | [0.001, 0.005] | Fitted |
| $\psi_2$ | 0.0013 | [0.001, 0.005] | Fitted |
| $\chi_{CM}$ | 1.2 | [1, 3] | Inferred from [10] |
| $\sigma_{CM}$ | 1.2 | [1, 3] | Assumed |
| $\delta_2$ | 0.0500 | [0.01, 0.08] | Fitted |
| $I_{CV}(0)$ | 135 | [100, 400] | Fitted |
| $I_{CVCm}(0)$ | 21 | [20, 50] | Fitted |
| $\mathcal{R}_{0c}$ | 1.3423 | | |

### 4.3 Discussion of results

Figure 3 presents the fitting of the model when the cumulative active cases were used to fit the model. The figure showed that the co-infection model (1) fitted well to the Lagos COVID-19 data (daily cumulative active cases).

As observed in Figure 4, the projection is made for the cumulative number of active cases using the developed model. It is observed that the cumulative active cases (including those with comorbidity) peaked at 130,000 cases in about 180 days (counting from March 16, 2020). Figure 5 presents the simulations of the cumulative number of active cases (including those with comorbidity) at different levels of susceptibility of comorbid susceptibles (0.2 ≤ *χ*_CM_ ≤ 1.2). It is observed that the peak value attained at different levels and reduces with decreasing value of the modification parameter *χ*_CM_. We notice that as preventive measures such as social distancing, use of face mask in public places are enforced to reduce the probability of infection by comorbid susceptibles, the infection peaks reduce stedily. Similar conclusions are reached when the cumulative number of active cases (without comorbidity) are simulated, as seen in Figure 6. The simulations of the cumulative number of active cases (including those with comorbidity), at different re-infection rates, depicted in Figure 7 shows the attainment of infections peaks around 200 days (counting from March 16, 2020), although infection peak reduces with decreasing re-infection of those who have recovered from a previous COVID-19 infection. Figure 8 is interpreted in a similar manner. It is also observed that, when the cumulative number of active cases are sumulated at different infectivity rates for co-infected individuals, the infecton curves behave in a similar trend, as when the modification parameter *χ*_CM_ are varied (See Figures 9 and 10).

**Figure 3:**
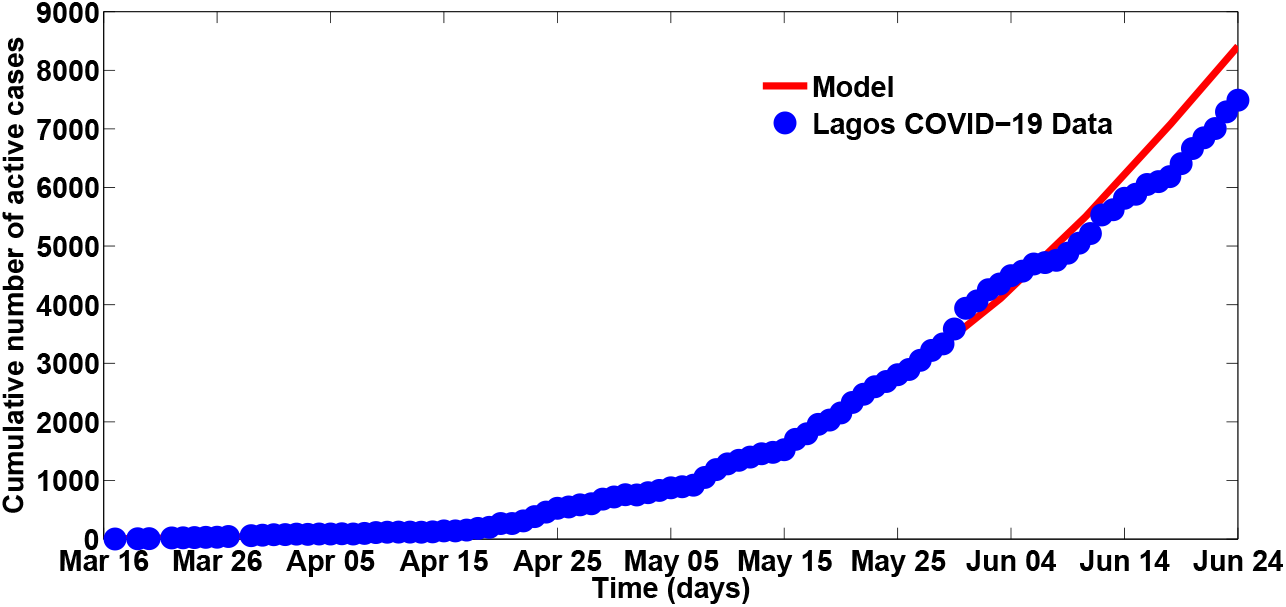
Fitting the cumulative number of active cases

**Figure 4:**
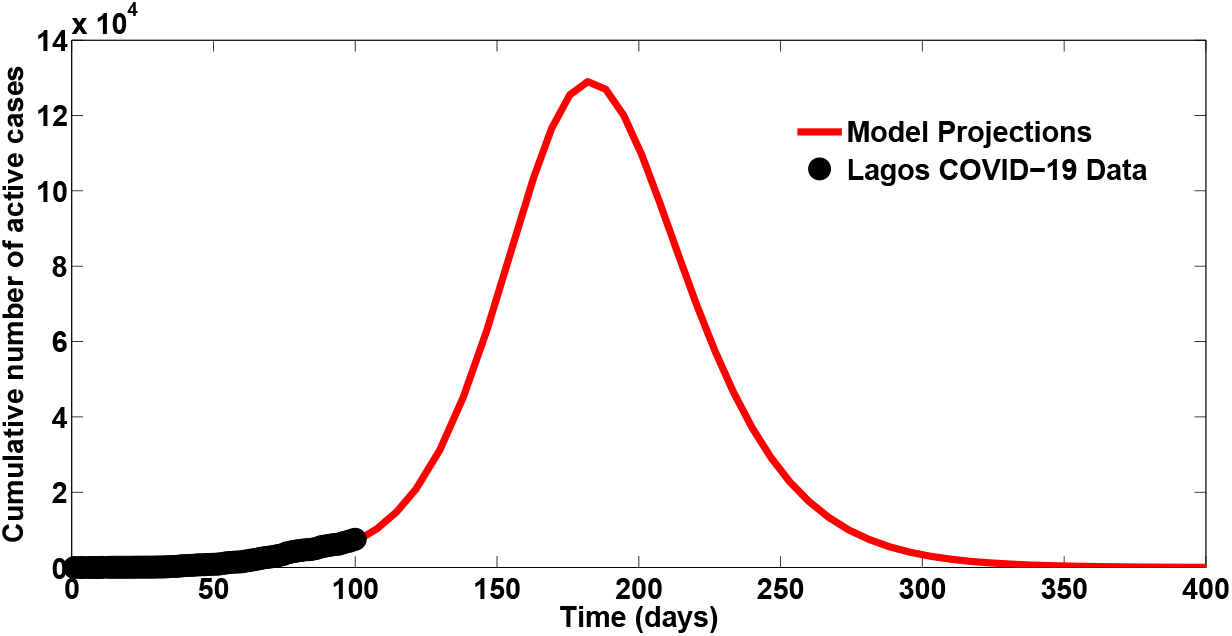
Projections for the cumulative number of active cases

**Figure 5:**
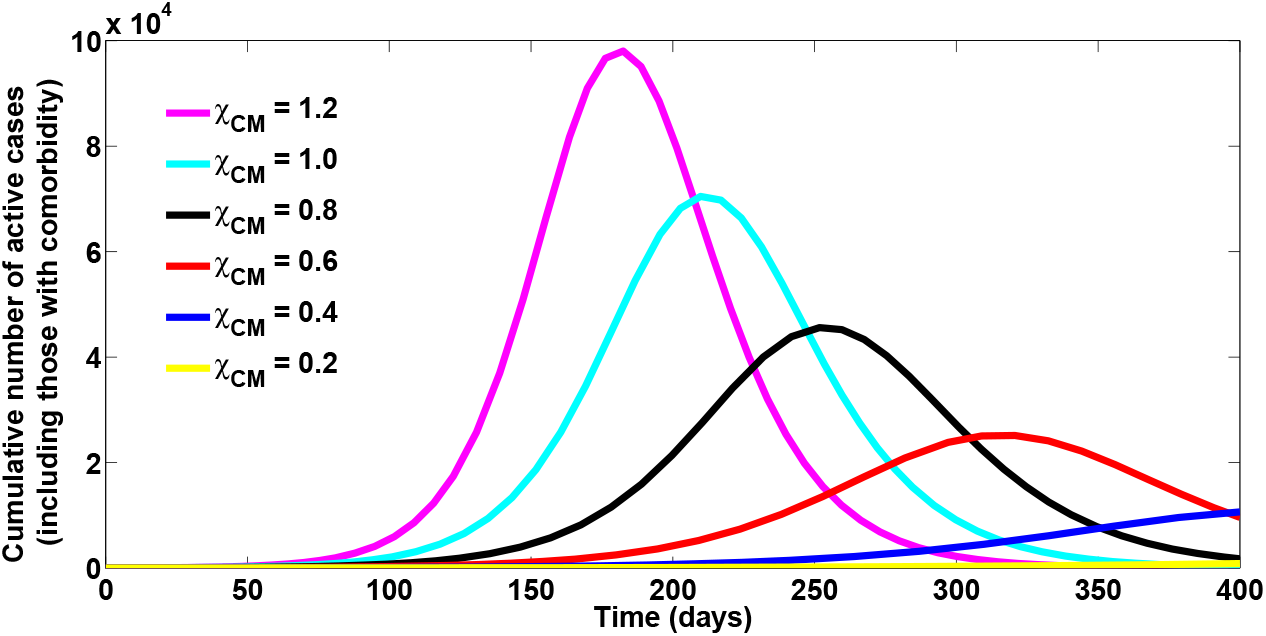
Simulations of the Cumulative number of active cases (including those with comorbidity): effect of *χ*_CM_

**Figure 6:**
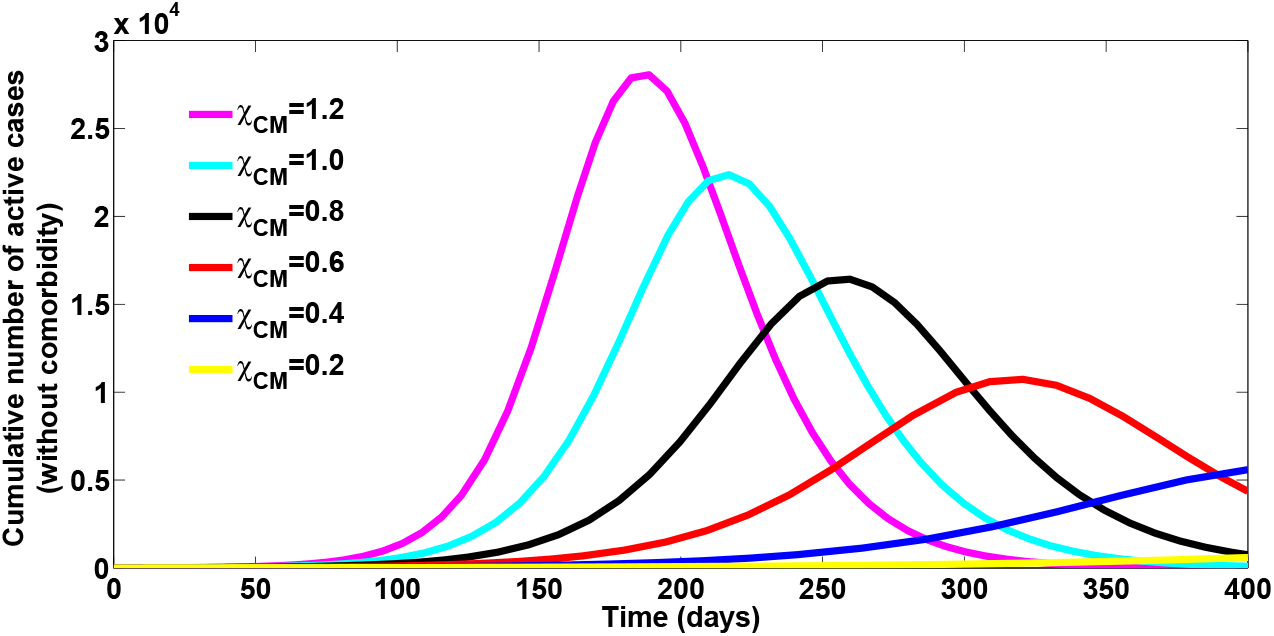
Simulations of the Cumulative number of active cases (without comorbidity): effect of *χ*_CM_

**Figure 7:**
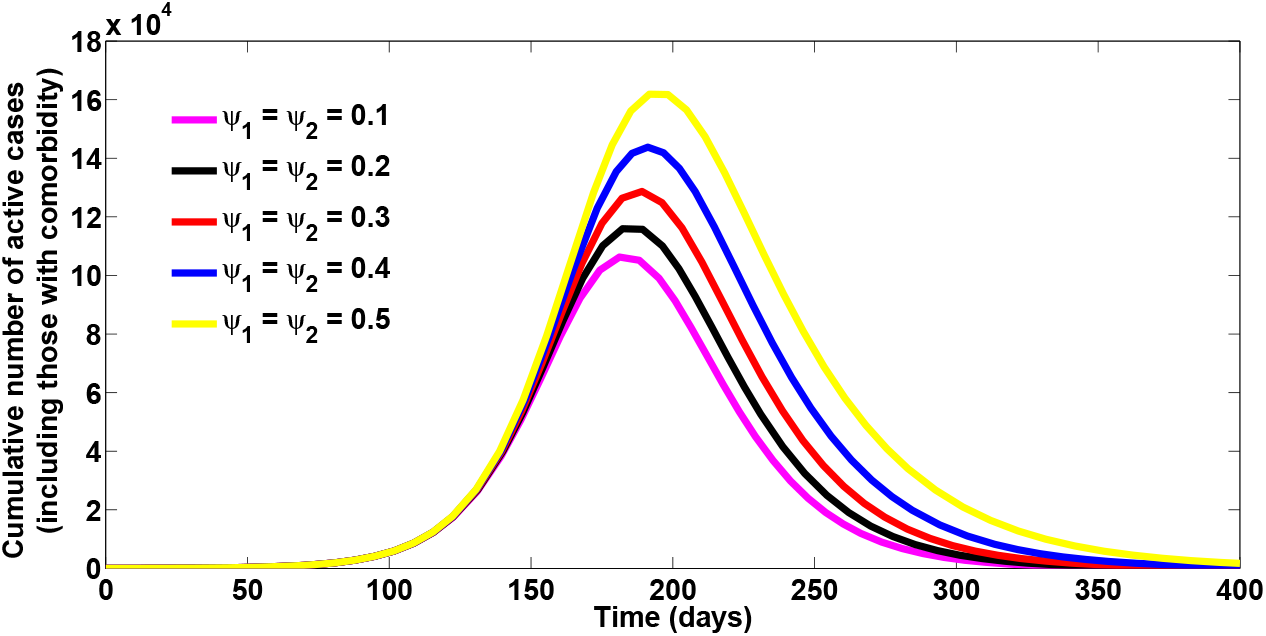
Simulations of the Cumulative number of active cases (including those with comorbidity): effect of *ψ*_1_ and *ψ*_2_

**Figure 8:**
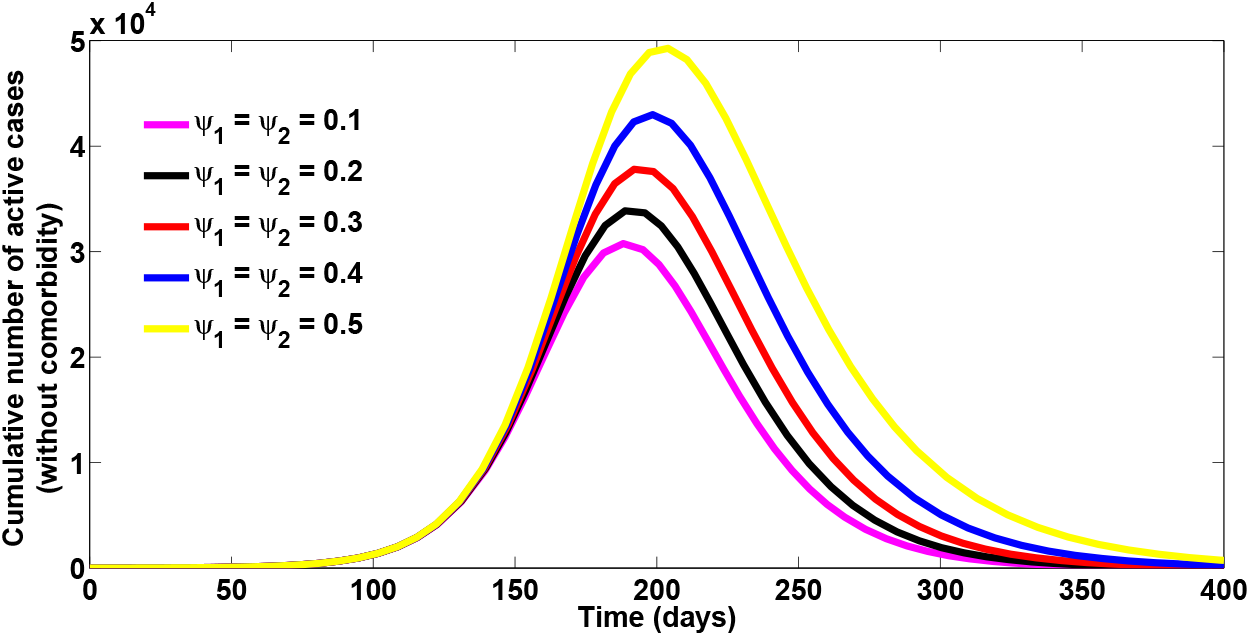
Simulations of the Cumulative number of active cases (without comorbidity): effect of *ψ*_1_ and *ψ*_2_

**Figure 9:**
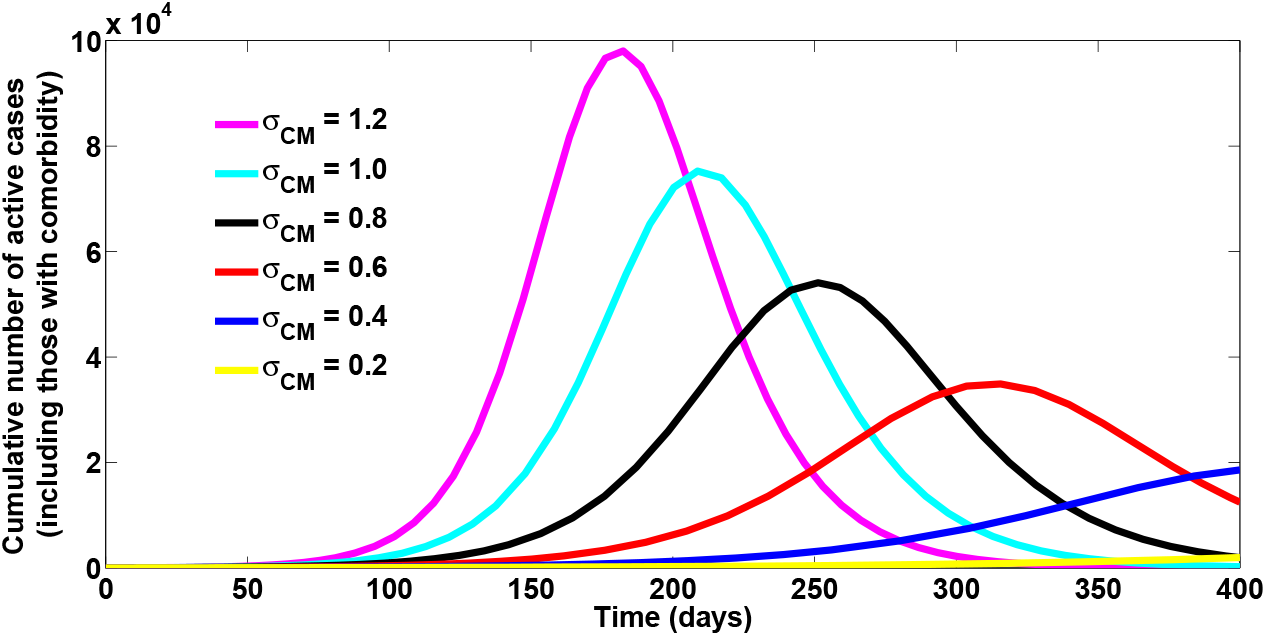
Simulations of the Cumulative number of active cases (including those with comorbidity): effect of *σ*_CM_

**Figure 10:**
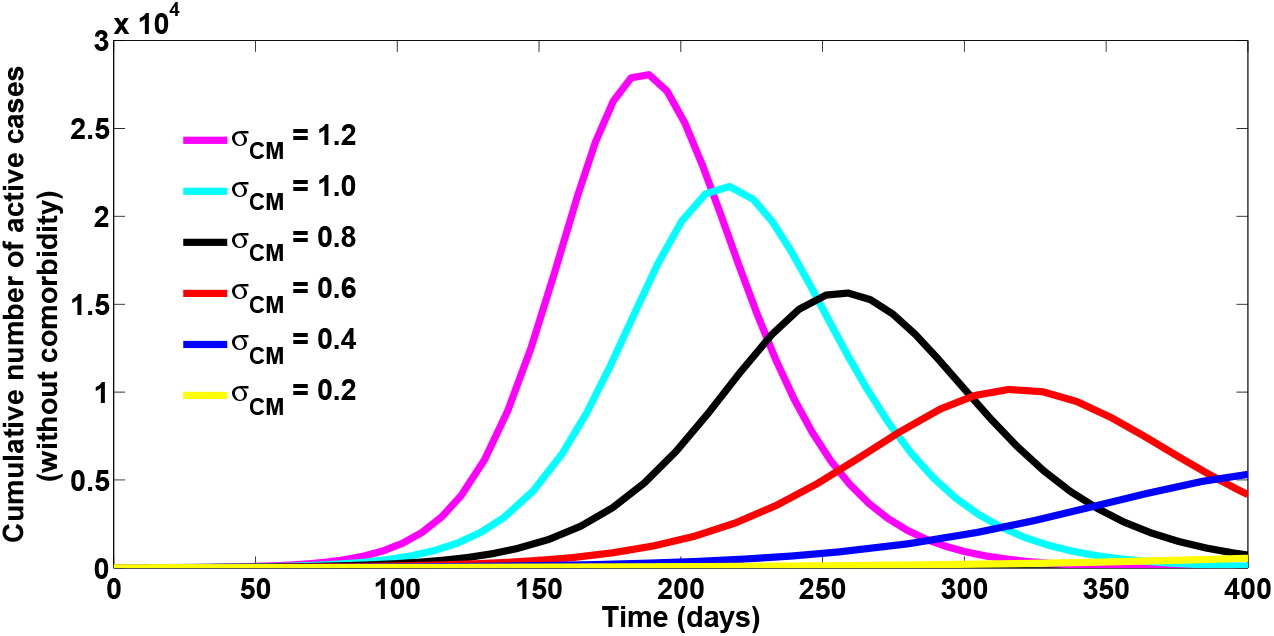
Simulations of the Cumulative number of active cases (without comorbidity): effect of *σ*_CM_

**Figure 11:**
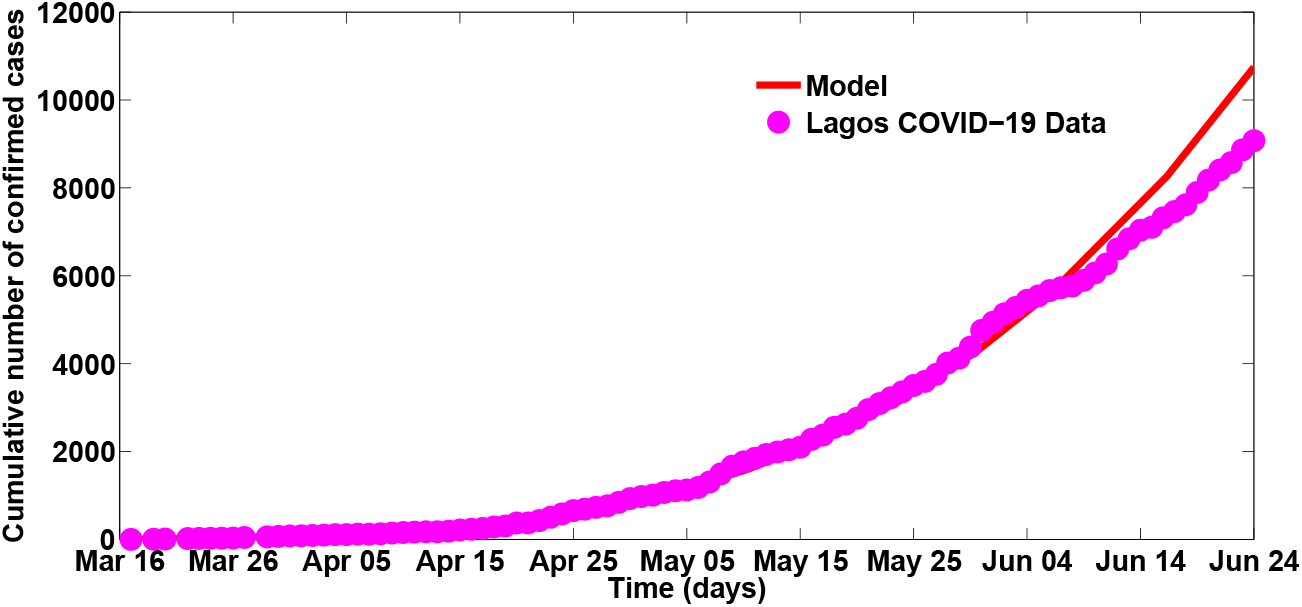
Fitting the cumulative number of confirmed cases

**Figure 12:**
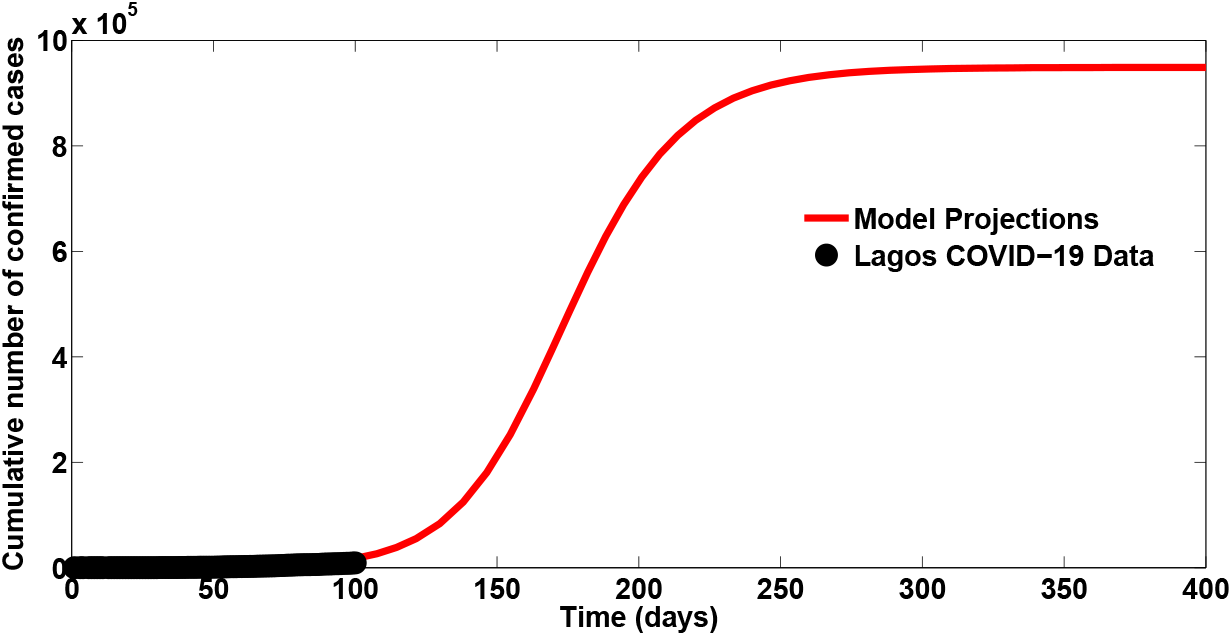
Projections for the cumulative number of confirmed cases

**Figure 13:**
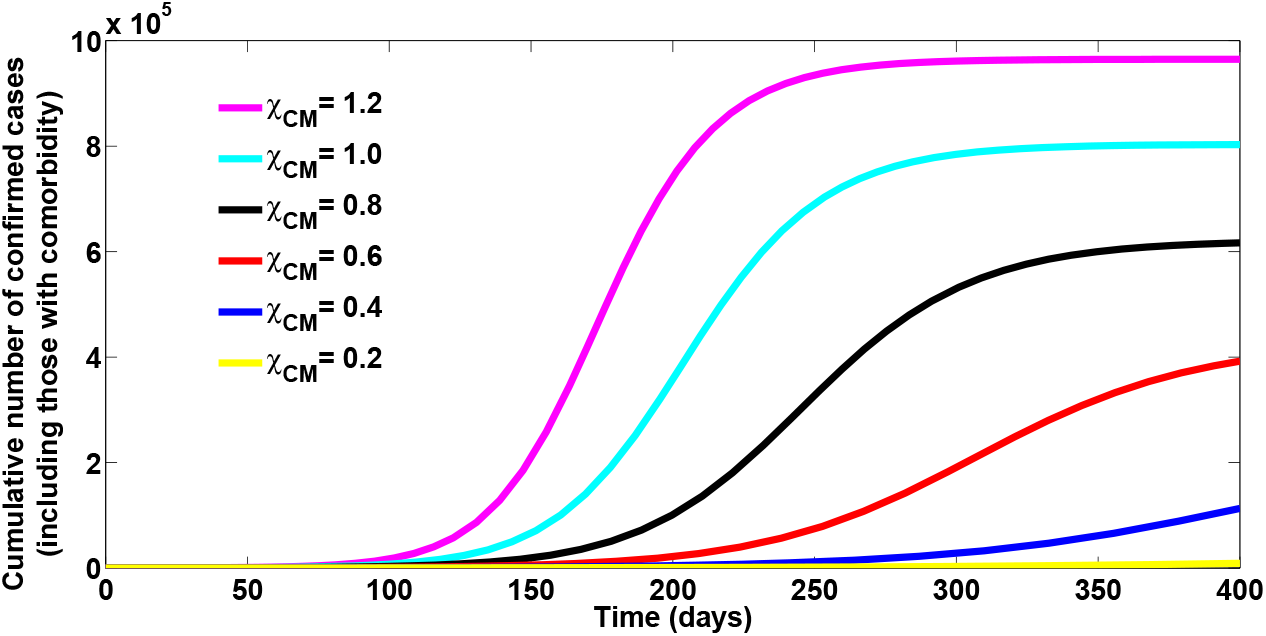
Simulations of the Cumulative number of confirmed cases (including those with comorbidity): effect of χ_CM_

**Figure 14:**
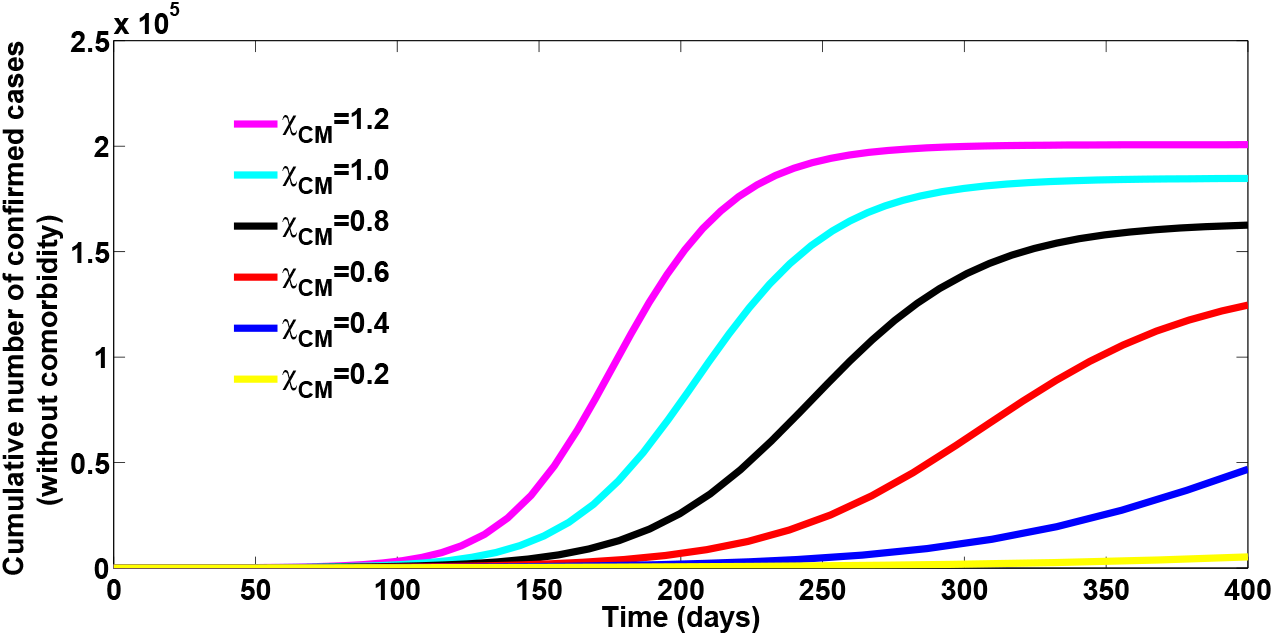
Simulations of the Cumulative number of confirmed cases (without comorbidity): effect of χ_CM_

**Figure 15:**
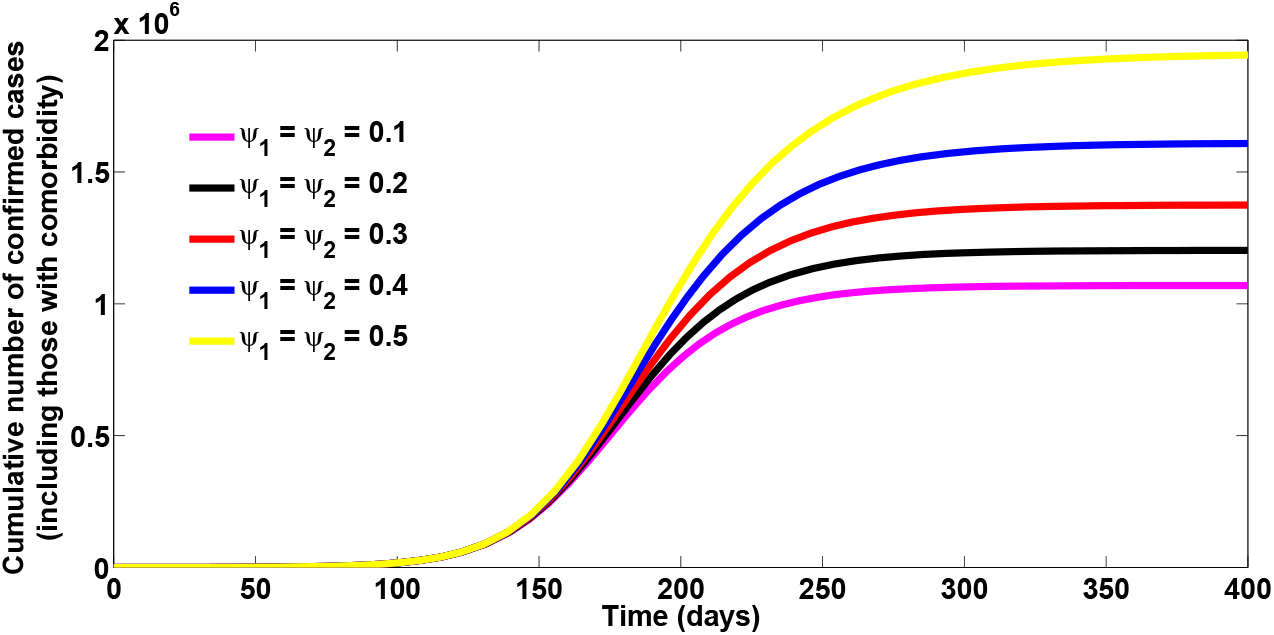
Simulations of the Cumulative number of confirmed cases (including those with comorbidity): effect of *ψ*_1_ and *ψ*_2_

**Figure 16:**
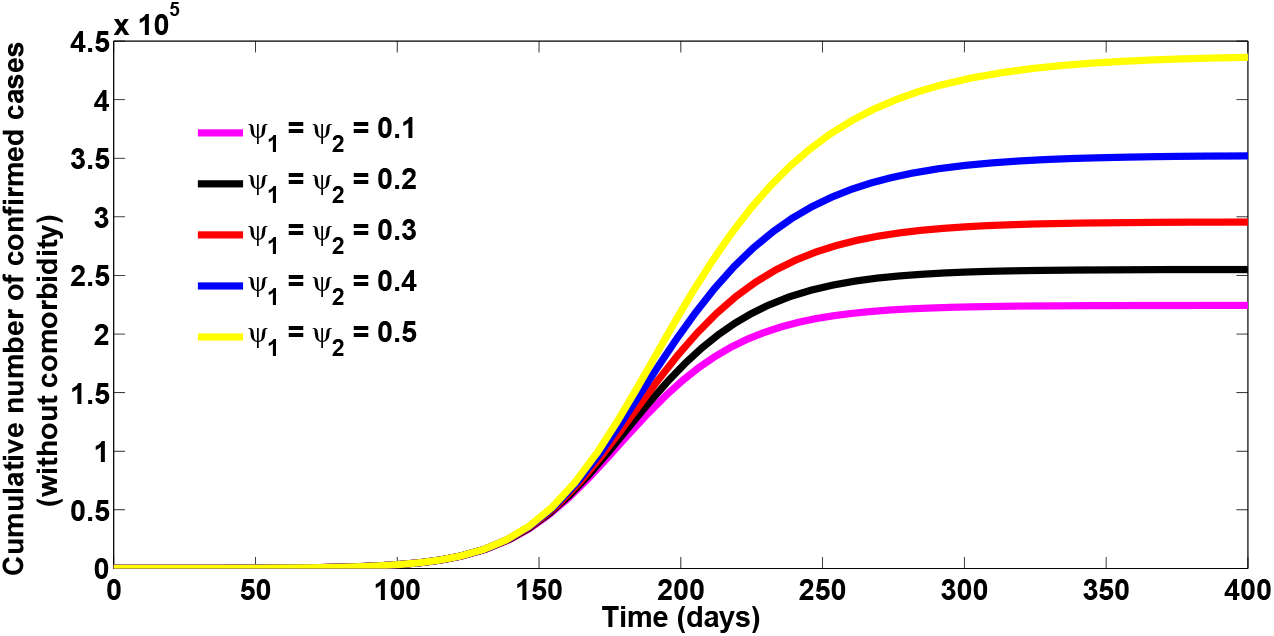
Simulations of the Cumulative number of confirmed cases (without comorbidity): effect of *ψ*_1_ and *ψ*_2_

**Figure 17:**
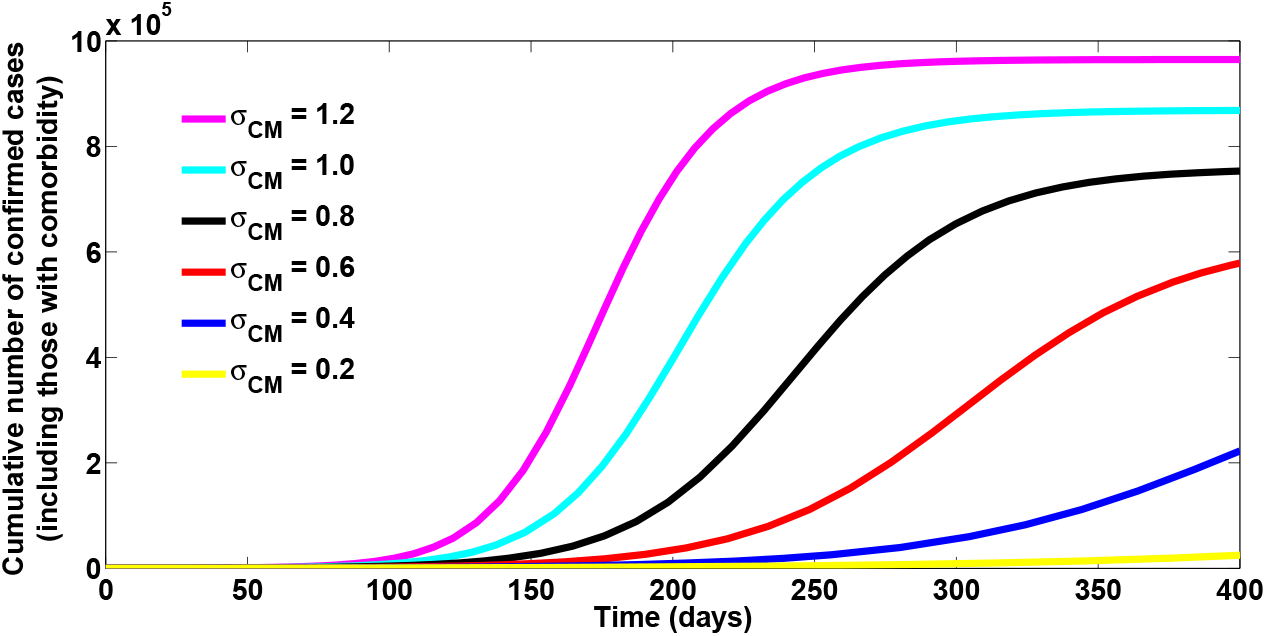
Simulations of the Cumulative number of confirmed cases (including those with comorbidity): effect of σ_CM_

**Figure 18:**
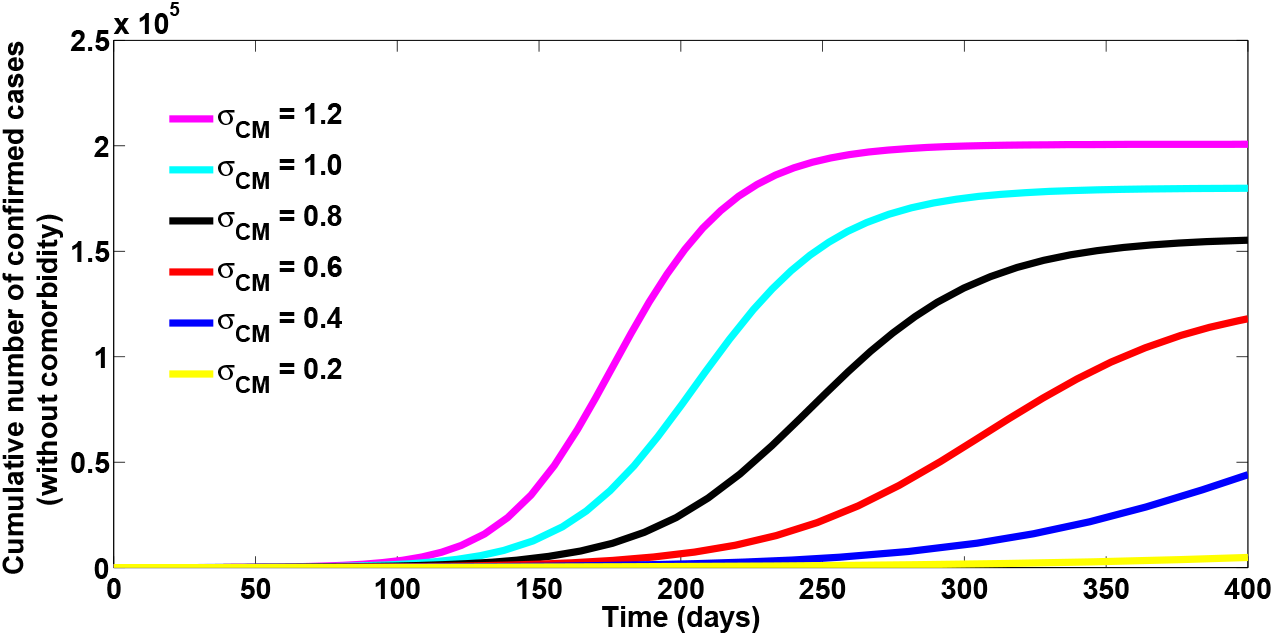
Simulations of the Cumulative number of confirmed cases (without comorbidity): effect of σ_CM_

Figure 11 depicts the fitting of the model when the cumulative confirmed cases were used to fit the model. The figure showed that the co-infection model (1) fitted well to the Lagos COVID-19 data (daily cumulative confirmed cases).

The projection of the cumulative confirmed cases, depicted in Figure 12, shows that in the absence of any preventive controls, the cumulative confirmed cases (including those co-infected with comorbidity) may reach 800,000 cases after 200 days (counting from the onset of infection in Lagos). As presented in Figure 13, the cumulative confirmed cases (including those with comorbidity) may gey up 900,000 cases after 200 days if the parameter accounting for hyper vulnerability of comorbid susceptibles is as high as 1.2 per day. The simulations of the cumulative confirmed cases (without comorbidity) may get up to 180,000 after 200 days, if the hyper susceptibility rate of comorbid susceptibles is as high as 1.2 per day (as depicted by Figure 14). However, this number decreases as efforts are strictly enforced to prevent COVID-19 infection among comorbid susceptibles, through the use of face-masks, observance of social distancing regulations. As illustrated in Figure 15, the cumulative confirmed cases (including those co-infected with comorbidity) may be as high as 1000,000 cases by the end of November, 2020 if the re-infection rates for COVID-19 is 0.1 per day. it may be worse than this if the re-infection rates increase higher. Similar conclusions are obtained when the cumulative confirmed cases (without comorbidity) are simulated at different re-infection rates, as depicted in Figure 16. When the cumulative confirmed cases with or without comorbidity are simulated at different infectivity levels for co-infected individuals (as presented in Figures 17 and 18), it is observed that the cumulative confirmed cases decreases with decreasing infectivity level for co-infected individuals.

The contour plot of the reproduction number 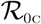 with detection rate for singly infected individuals (*η*_1_) and detection rate for co-infected individuals (*η*_2_), shown in Figure 19, reveal that if the detection rate for infected individuals co-infected with comorbidity can be increased to about 0.7 per day and the detection rate for infected individuals (without comorbidites) kept at 0.3 per day, then the reproduction number 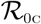 can be brought below unity and COVID-19 infection eliminated from the population. The contour plot of the reproduction number, 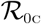, as a function of detection rate for singly infected individuals (*η*_1_) and the modification parameter for high suscptibility of comorbid susceptibles, depicted in Figure 20 shows that if policies are strictly put in place to step down the probability of COVID-19 infection by comorbid susceptibles to as low as 0.4 per day and step up the detection rate for singly infected individuals to 0.7 per day, then the reproduction number can be brought very low below one, and COVID-19 infection eliminated from the population.

**Figure 19:**
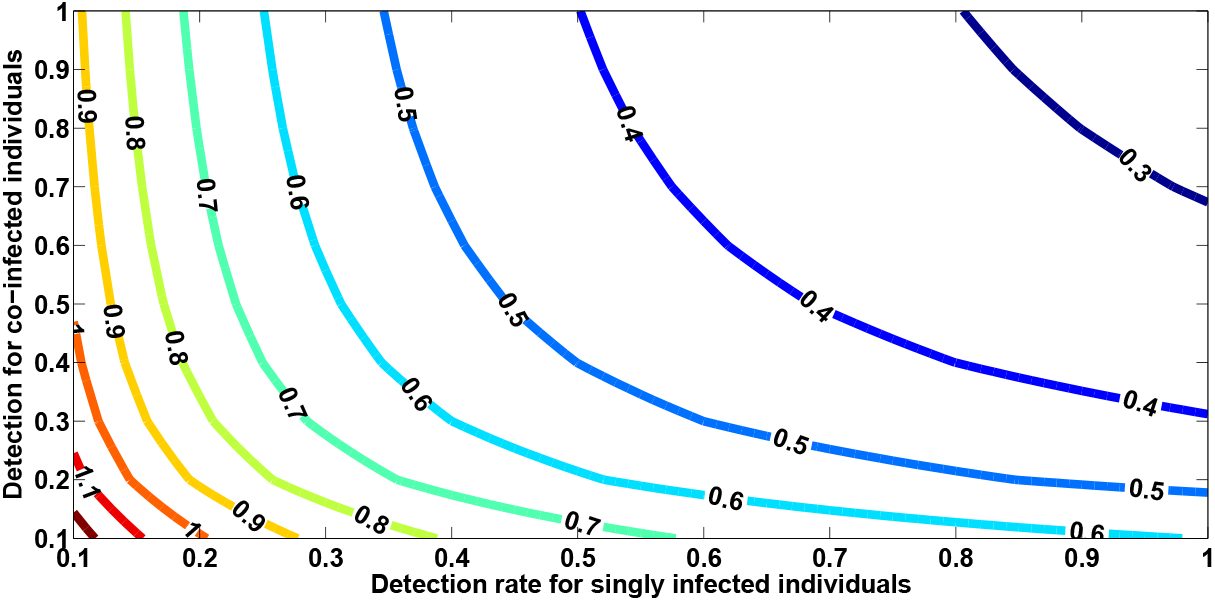
A contour plot of 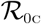 as a function of detection rate for singly infected individuals (*η*_1_) and detection rate for co-infected individuals (*η*_2_)

**Figure 20:**
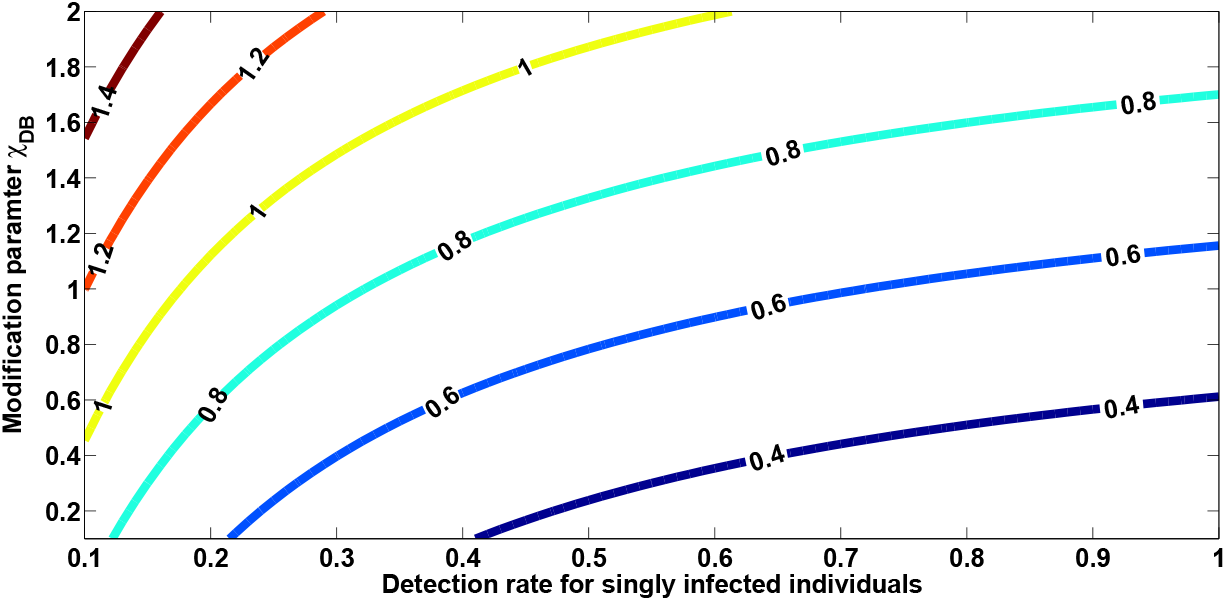
A contour plot of 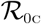 as a function of detection rate for singly infected individuals (*η*_1_) and the modification parameter for high susceptibility of those with comorbidity (χ_CM_)

## 5 Analysis of the optimal control model

The Pontryagin’s Maximum Principle shall be applied, in this section, to determine the necessary conditions for the optimal control of the model. We incorporate time dependent controls into the model (1) to determine the optimal strategy for controlling the spread of COVID-19. Thus, we have,

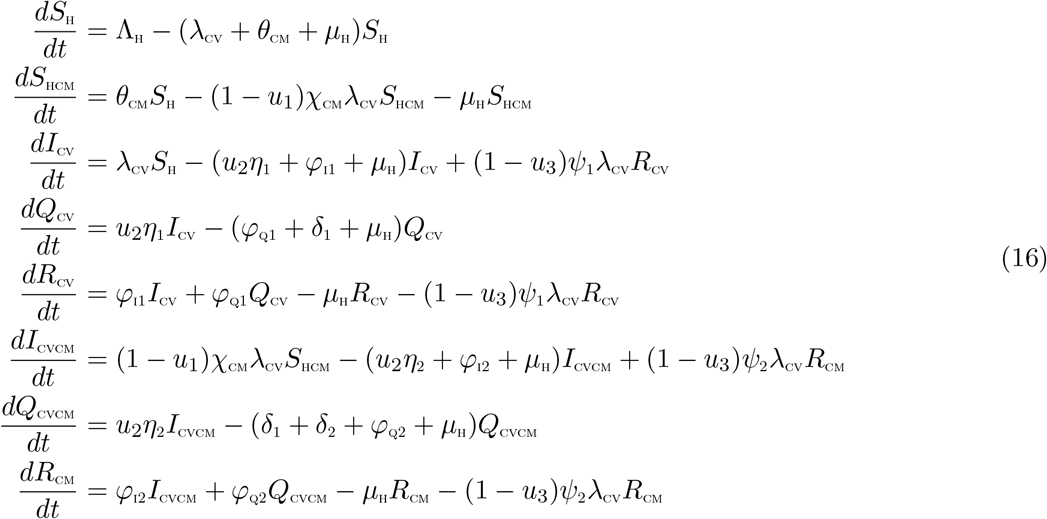

subject to the initial conditions

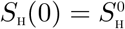, 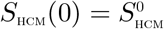, 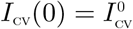, 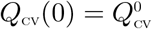, 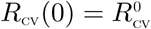, 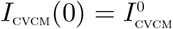, 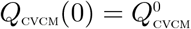, 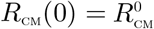

with

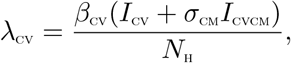

The control functions, *u*_1_(*t*), *u*_2_(*t*), and *u*_3_(*t*) are bounded, Lebesgue integrable functions. The control *u*_1_(*t*) represents the efforts geered towards preventing incident COVID-19 infections by comorbid suceptible humans. The control *u*_2_(*t*) is the effort aimed at intensifying detection of COVID-19 cases. Efforts aimed at preventing re-infection by those who have fully recovered from COVID-19 is represented by *u*_3_(*t*). The control *u*_1_ satisfies 0 ≤ *u*_1_ ≤ 0.9, the controls u_2_ and u_3_ satisfy 0 < *u*_2_, *u*_3_ ≤ 1. The optimal control system examines scenarios where the number of infectious cases and the cost of implementing the controls *u*_1_(*t*), *u*_2_(*t*), and *u*_3_(*t*) are minimized subject to the state system (16). For this, we consider the objective functional

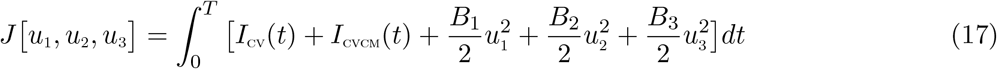

*T* is the final time. We seek to find an optimal control, 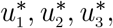 such that

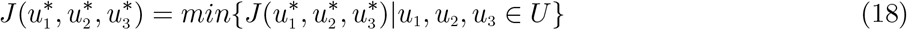

where 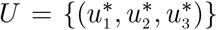 such that 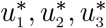 are measurable with 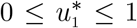, 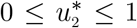, 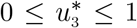, for *t* ∈ [0, *T*] is the control set. The Pontryagin’s Maximum Principle [37] gives the necessary conditions which an optimal control pair must satisfy. This principle transforms (16), (17) and (18) into a problem of minimizing a Hamiltonian, 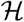, pointwisely with regards to the control functions, *u*_1_, *u*_2_, *u*_3_:

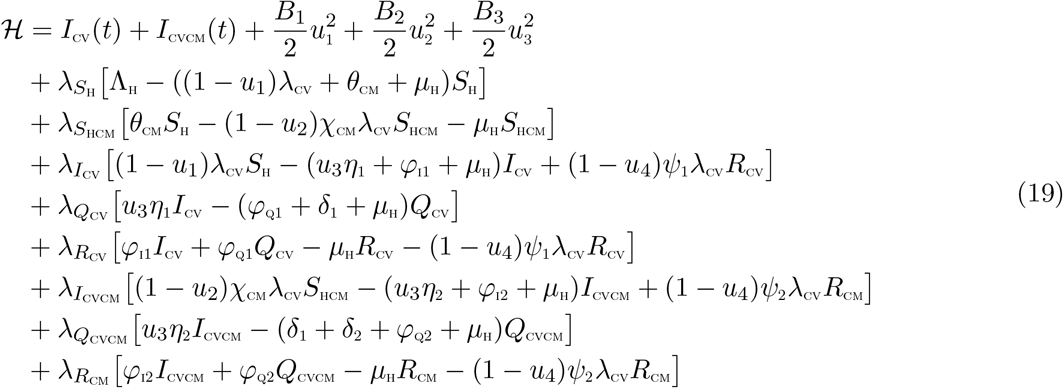

### Theorem 5.1

*For an optimal control set u*_1_, *u*_2_, *u*_3_ *that minimizes J over U, there are adjoint variables, λ*_1_, *λ*_2_, …, *λ*_8_ *satisfying*

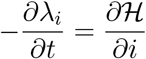

*and with transversality conditions*

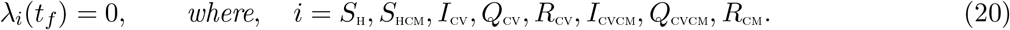

*Furthermore*,

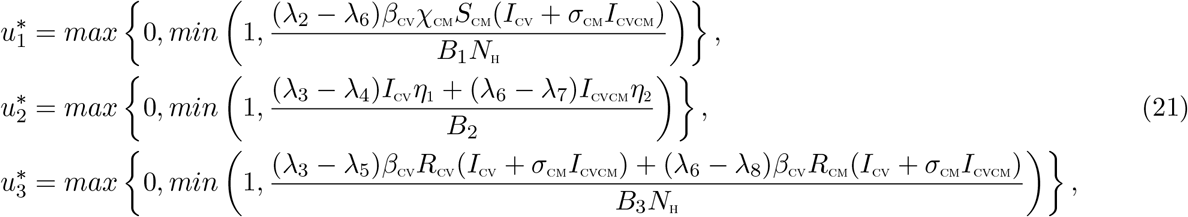

#### Proof of Theorem 5.1

Suppose 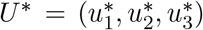 is an optimal control and 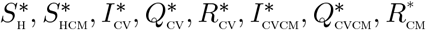 are the corresponding state solutions. Applying the Pontryagin’s Maximum Principle [37], there exist adjoint variables satisfying:

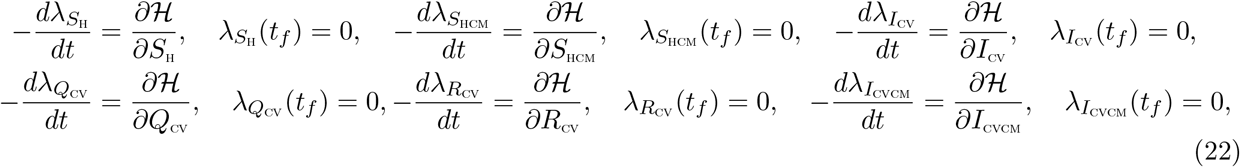

with transversality conditions;

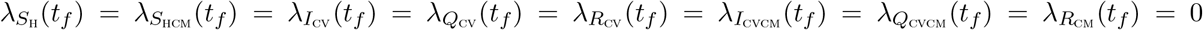 We can determine the behaviour of the control by differentiating the Hamiltonian, 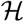 with respect to the controls(*u*_1_, *u*_2_, *u*_3_) at *t*. On the interior of the control set, where 0 < *u_j_* < 1 for all (*j* = 1, 2, 3), we obtain

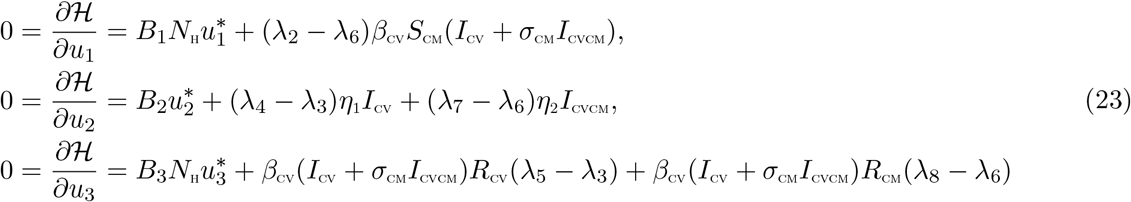

Therefore, we have that [25]

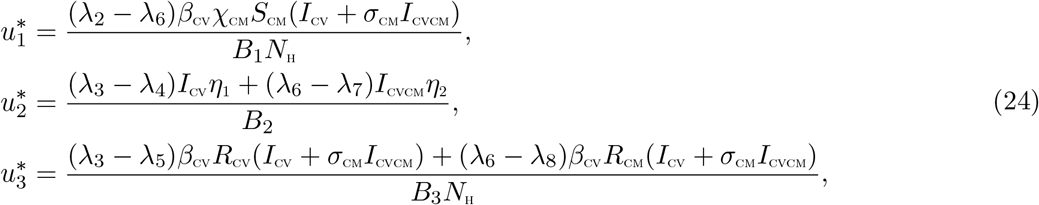

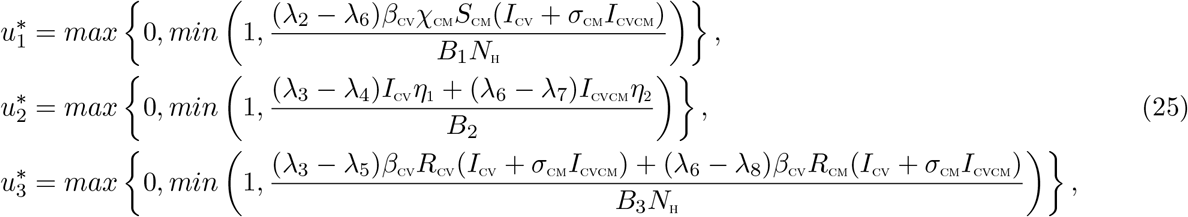

**Figure 21:**
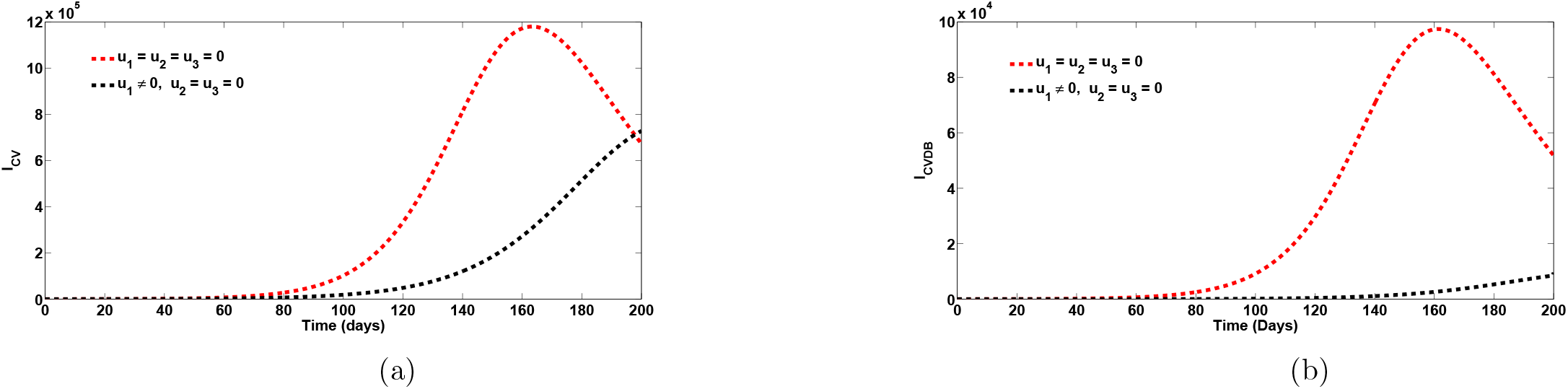
Plots of the total number of COVID-19 infected individuals when strategy A is implemented (*u*_1_ ≠ 0). Here, *β*_CV_ = 0:2001; *ψ*_1_ = *ψ*_2_ = 0:4. All other parameters as in Table 1

**Figure 22:**
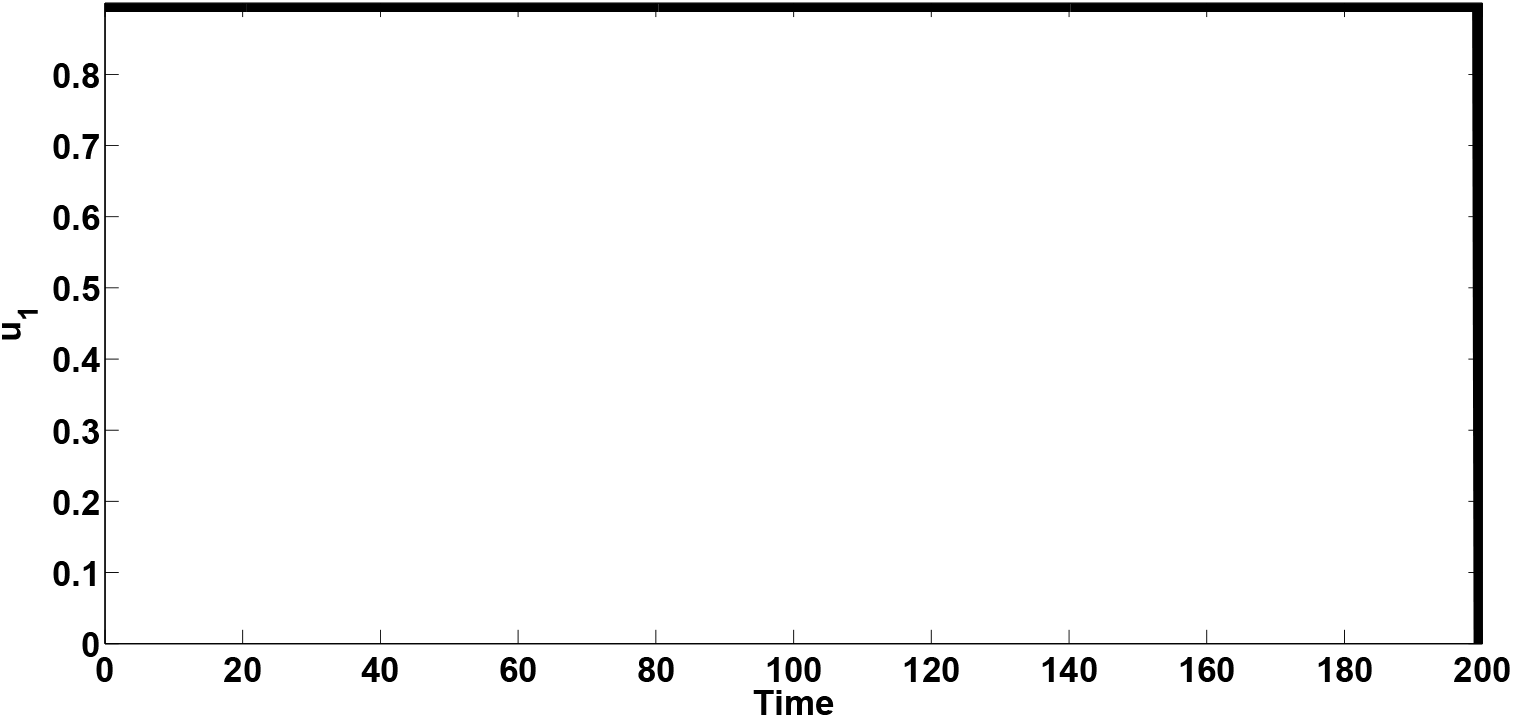
Control profile for the effect of the control *u*_1_ on the dynamics of the model (1). Here, *β*_CV_ = 0.2001, *ψ*_1_ = *ψ*_2_ = 0.4. All other parameters as in Table 1

### 5.1 Strategy A: COVID-19 prevention among Comorbid susceptibles (*u*_1_ ≠ 0)

Simulations of the optimal control system when the strategy that prevents COVID-19 among comorbid susceptibles (*u*_1_ ≠ 0) is implemented, are presented in Figure 21. It is seen that when this intervention strategy is implemented, there is a significant decrease in the total number of individuals singly infected with COVID-19 (Figure 21(a)) and those co-infected with COVID-19 and comorbidity (Figure 21 (b)). Specifically, this strategy averts 908,500 new cases of COVID-19 and also prevents 94,747 new cases of co-infection of COVID-19 and comorbidity after 160 days. The control profile for this control strategy given in Figure 22 shows that control *u*_1_ is at its peak throughout the entire simulation period.

### 5.2 Strategy B: Case detection control (*u*_2_ ≠ 0)

Simulations of the optimal control system when the case detection control (*u*_2_ ≠ 0) is implemented, are presented in Figure 23. It is seen that when this intervention strategy is implemented, there is a significant drop in the total number of individuals singly infected with COVID-19 and the co-infection cases. To be specific, this strategy averts 657,100 new COVID-19 cases after 160 days. The strategy also averts 67,230 new co-infection cases after 160 days. The control profile for this intervention control given in Figure 24 also shows that control *u*_2_ is at its maximum value for the entire period of the simulation.

### 5.3 Strategy C: Control against COVID-19 re-infection (*u*_3_ ≠ 0)

Simulations of the optimal control system when control against COVID-19 re-infection (*u*_3_ ≠ 0) is implemented, are presented in Figure 25. It is observed that when this intervention strategy is applied, there is a significant reduction in the total number of individuals singly infected with COVID-19 and those co-infected with comorbidity. As a matter of fact, this strategy averts 431,300 new COVID-19 cases and 34,620 co-infection new cases. The control profile for this intervention control given in Figure 26 shows that control *u*_3_ rises steadily to its maximum value in about 50 days and maintains this peak for the entire period of the simulation.

**Figure 23:**
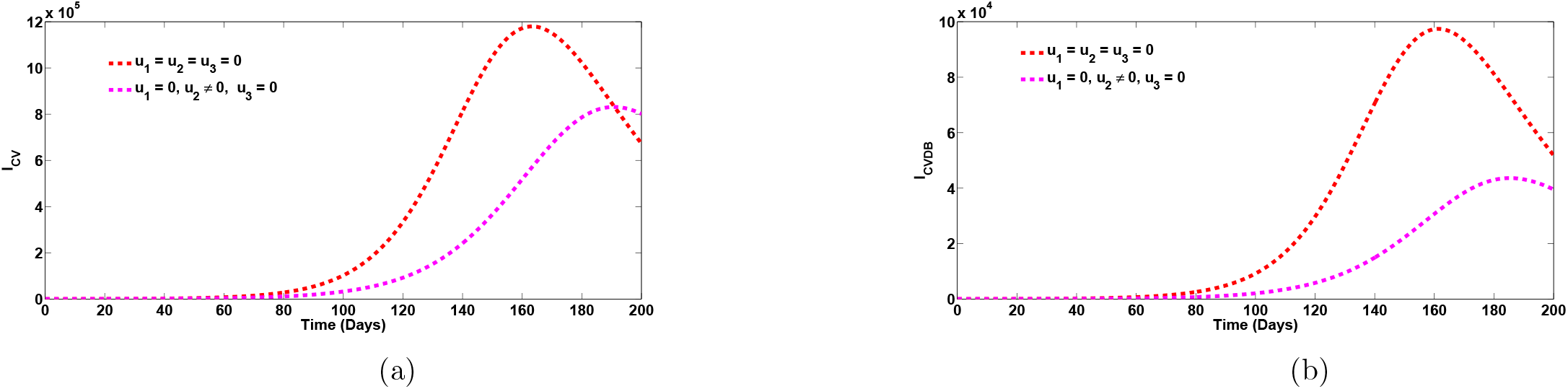
Plots of the total number of COVID-19 infected individuals when strategy B is implemented (*u*_2_ ≠ 0). Here, *β*_CV_ = 0.2001, *ψ*_1_ = *ψ*_2_ = 0.4. All other parameters as in Table 1

**Figure 24:**
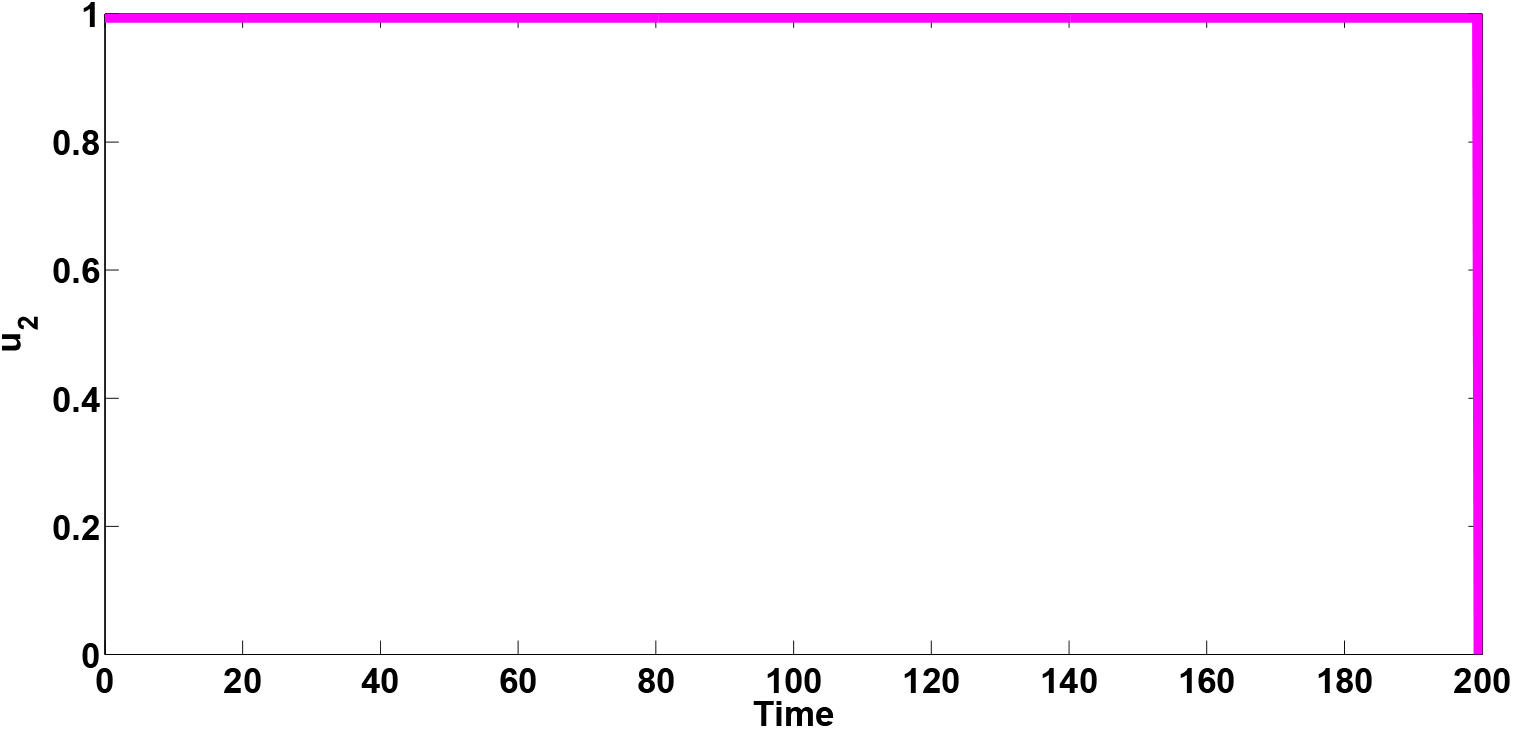
Control profile for the effect of the control *u*_2_ on the dynamics of the model (1). Here, *β*_CV_ = 0.2001, *ψ*_1_ = *ψ*_2_ = 0.4. All other parameters as in Table 1

**Figure 25:**
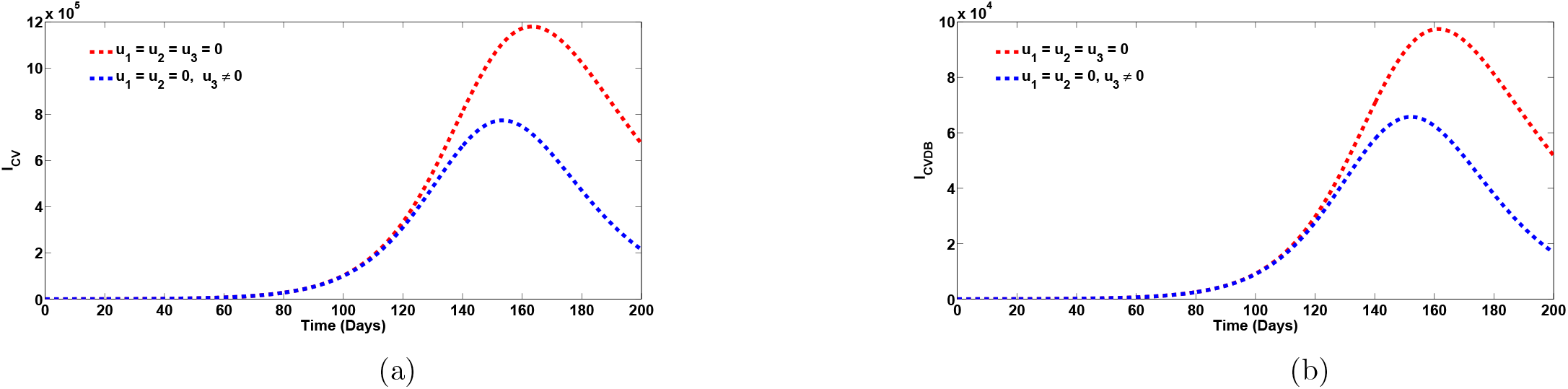
Plots of the total number of COVID-19 infected individuals when strategy C is implemented (*u*_3_ ≠ 0). Here, *β*_CV_ = 0.2001, *ψ*_1_ = *ψ*_2_ = 0.4. All other parameters as in Table 1

**Figure 26:**
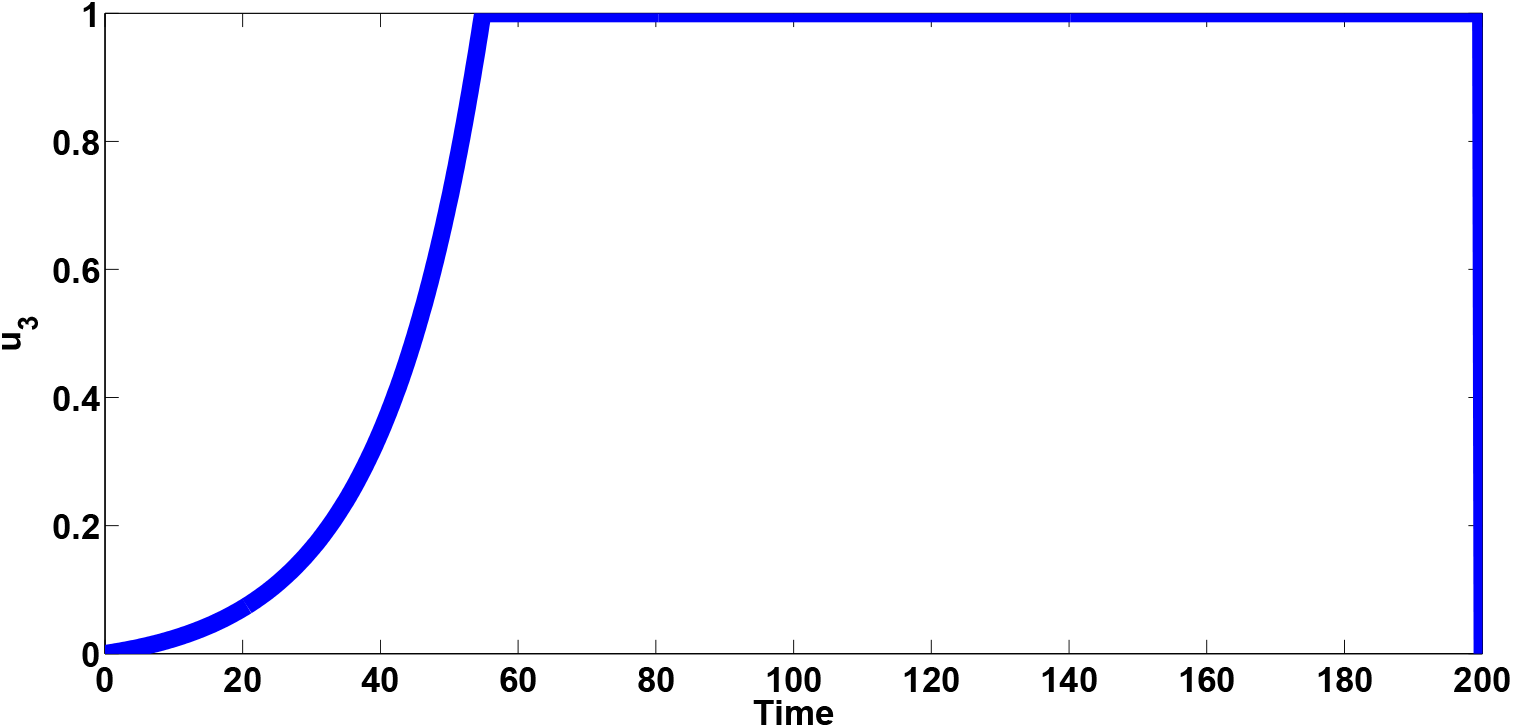
Control profile for the effect of the control *u*_3_ on the dynamics of the model (1). Here, *β*_CV_ = 0.2001, *ψ*_1_ = *ψ*_2_ = 0.4. All other parameters as in Table 1

### 5.4 Cost-effectiveness analysis

The cost-effectiveness analysis is used to evaluate the health interventions related benefits in order to justify the costs of the strategies [7]. This is obtained by comparing the differences among the health outcomes and costs of those interventions; achieved by computing the incremental cost-effectiveness ratio (ICER), which is defined as the cost per health outcome.

We calculated the total number of cases averted and the total cost of the strategies applied in Table 4. The total number of cases prevented is obtained by calculating the total number of individuals when controls are implemented and the total number when control is not administered. In a similar manner, we apply the cost functions 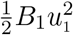, 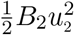, 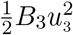, over time, to compute the total cost for the various strategies that were carried out. We assume the weight constants *B*_1_ = 400, *B*_3_ = 500 while *B*_2_ = 700. We assumed here that, the cost of implementing the case detection control is much higher compared to the cost of implementing the preventive controls (which are mainly non-pharmaceutical). The cost-effectiveness of strategy C (control against re-infection by those who have recovered from a previous COVID-19 infection) and strategy B (case detection control) are now compared.

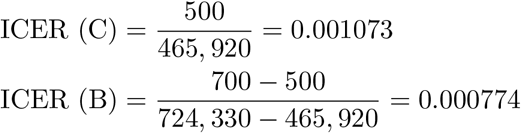

**Table 4:**
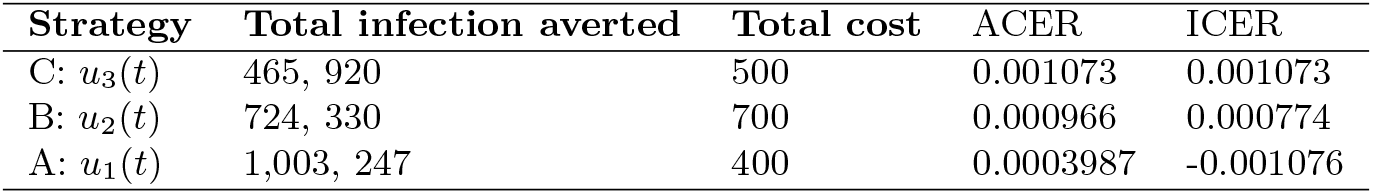
Increasing order of the total infection averted due to the control strategies

**Table 5:** Increasing order of the total infection averted due to the control strategies

| Strategy | Total infection averted | Total cost | ACER | ICER |
| --- | --- | --- | --- | --- |
| B: $u_2(t)$ | 724, 330 | 700 | 0.000966 | 0.000966 |
| A: $u_1(t)$ | 1,003, 247 | 400 | 0.0003987 | -0.001076 |

From ICER (C) and ICER(B), we observe a cost saving of 0.000774 observed for strategy B over strategy C. This implies that strategy C strongly dominated strategy B, showing that strategy C is more costly and less effective compared to strategy B. Therefore, strategy C is removed from subsequent ICER computations, shown in Table 5. We shall now compare strategies B and A. Comparing strategy B (case detection control) and strategy A (control against COVID-19 infection by comorbid susceptibles), we observe that ICER (B) is greater than ICER (A), showing that strategy B strongly dominated strategy A and is more expensive and less effective compared to strategy A. Consequently, strategy A (the strategy that prevents COVID-19 infection by comorbid susceptibles) has the least ICER and is the most cost-effective of all the control strategies for the prevention of COVID-19. This also conforms with the results obtained from ACER method in Table 4 that strategy A is the most cost-effective strategy. It is also worthy of note to observe that this strategy averts more new COVID-19 cases than any other control strategy implemented.

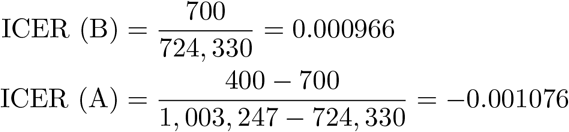

## 6 Conclusion

In this work, we developed and analyzed a mathematical model for the dynamics of COVID-19 infection in order to assess the impacts of prior comorbidity on COVID-19 complications and COVID-19 re-infection. The model was simulated using data relevant to the dynamics of the diseases in Lagos, Nigeria, making predictions for the attainment of peak periods in the presence or absence of comorbidity. The model was shown to undergo the phenomenon of backward bifurcation which was caused by the parameter accounting for increased susceptibility to COVID-19 infection by comorbid susceptibles as well as the rate of re-infection by those who have recovered from a previous COVID-19 infection. The epidemiological interpretation of this phenomenon is that if recovery from COVID-19 does not confer lifelong immunity and efforts are not put in place to prevent COVID-19 infections by comorbid susceptible individuals, then the control of COVID-19 becomes difficult in the population, even when the associated reproduction number 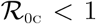. Hence, it is recomended that efforts should be made by those in authority and policy makers to recommend preventive measures to curb COVID-19 infection by comorbid susceptibles and re-infection with COVID-19 so as to bring the burden of COVID-19 infection very low at the community level.

Sensivity analysis of the model when the population of individuals co-infected with COVID-19 and comorbidity is used as response function revealed that the top ranked parameters that drive the dynamics of the co-infection model are the effective contact rate for COVID-19 transmission, *β*_CV_, the parameter accounting for increased sucseptibility to COVID-19 by comorbid susceptibles, *χ*_CM_, the comorbidity development rate, *θ*_CM_, the detection rate for singly infected and co-infected individuals, *η*_1_ and *η*_2_, as well as the recovery rate from COVID-19 for co-infected individuals, *φ*_I2_. Simulations of the model reveal that the cumulative confirmed cases (without comorbidity) may get up to 180,000 after 200 days, if the hyper susceptibility rate of comorbid susceptibles is as high as 1.2 per day. Moreover, the cumulative confirmed cases (including those co-infected with comorbidity) may be as high as 1000,000 cases by the end of November, 2020 if the re-infection rates for COVID-19 is 0.1 per day. It may be worse than this if the re-infection rates increase higher. In addition, if policies were strictly put in place to step down the probability of COVID-19 infection by comorbid susceptibles to as low as 0.4 per day and the detection rate for singly infected individuals stepped up to 0.7 per day, then the reproduction number could be brought very low below unity, and COVID-19 infection eliminated from the population. Optimal control and cost effectiveness analysis of the model reveals that the strategy that prevents COVID-19 infection by comorbid susceptibles has the least ICER and is the most cost-effective of all the control strategies for the prevention of COVID-19.

## Data Availability

Data available on https://covid19.ncdc.gov.ng

